# GlucoseGo: A Simple, User-Friendly, Machine Learning-Derived Tool for Predicting Exercise-Related Hypoglycaemia Risk in Type 1 Diabetes

**DOI:** 10.1101/2025.04.24.25326242

**Authors:** Catherine CL Russon, Michael MJ Allen, Richard RM Pulsford, Neil N Vaughan, Emma E Cockcroft, John J Pemberton, Anne Marie A Frohock, Robert RC Andrews

## Abstract

**Aims/hypothesis:** This study aims to develop an accessible, machine learning-derived tool for people with type 1 diabetes that predicts hypoglycaemia risk at the start of exercise, facilitating quick, clear risk assessment that can directly support safer exercise habits.

**Methods:** We integrated data from four diverse studies encompassing 16,477 exercise sessions from 834 participants aged 12-80, using various insulin delivery methods. The XGBoost algorithm was used to develop a comprehensive and simplified model to predict hypoglycaemia during exercise, determined by continuous glucose monitor readings below 3.9 mmol/L (70 mg/dL).

**Results:** The comprehensive model demonstrated a mean ROC AUC of 0.89, while the simplified model, relying solely on glucose levels at the start of exercise, duration of exercise and glucose rate of change arrows, achieved an ROC AUC of 0.87. This model was shown to be effective for any type of exercise and for people on a variety of insulin delivery devices. This simplified model was then translated, through collaborative efforts with type 1 diabetes participants, into “GlucoseGo,” a user-friendly, traffic-light heatmap that visually demonstrates risk of hypoglycaemia during exercise based on these three variables.

Conclusions/interpretation

The GlucoseGo heatmap offers a simple, readily available tool for predicting hypoglycaemia risk at the onset of exercise. This advancement empowers users to manage their exercise routines more safely, with potential to reduce hypoglycaemia incidents and enhancing exercise engagement among the type 1 diabetes population.

**Research in context:** *What is already known about this subject?:*

- Exercise is crucial for managing type 1 diabetes, yet adherence to recommended guidelines is low.
- Exercise-induced hypoglycaemia is a major barrier to exercise for those with type 1 diabetes.
- Existing machine learning models for predicting hypoglycaemia during exercise often require complex data inputs, limiting their practical use.

*What is the key question?:*

- Can a machine learning model using minimal data effectively predict exercise-induced hypoglycaemia in type 1 diabetes?

*What are the new findings?:*

- We developed a simplified machine learning model using only three variables; starting glucose levels, glucose rate of change arrows and exercise duration - that nearly matches the performance of more complex models, with an ROC AUC of 0.87 versus 0.89.
- This model was transformed into “GlucoseGo,” user-friendly heatmaps, designed collaboratively with individuals with type 1 diabetes, that visually indicate exercise-induced hypoglycaemia risk.
- Subgroup analyses show consistently good performance in predicting hypoglycaemia risk across diverse patient profiles and exercise types, validating its broad applicability.

*How might this impact clinical practice in the foreseeable future?:* GlucoseGo offers a practical tool for safely managing exercise, potentially reducing hypoglycaemic incidents and increasing exercise participation among those with type 1 diabetes.

## Introduction

Exercise is crucial in the management of type 1 diabetes, providing significant benefits including improved glycaemic control, enhanced cardiovascular health, improved psychological wellbeing and reduced long term complications (Bohn et al., 2015; Codella et al., 2017; Katz et al., 2015; R. G. Miller et al., 2016; Pierre-Louis et al., 2014). Yet adherence to exercise guidelines remains markedly low, with ∼60% not achieving recommendations (Bohn et al., 2015; Finn et al., 2022; Keshawarz et al., 2018). There are many factors that contribute to this, but the strongest barrier to exercise is repeatedly reported to be fear of exercise-induced hypoglycaemia (Brazeau et al., 2008; Brennan et al., 2021; Cigrovski Berković et al., 2021; Finn et al., 2022; Lascar et al., 2014; McCarthy et al., 2016). Hypoglycaemia not only poses immediate health risks but also has a psychological impact that can discourage regular exercise, thereby compounding the long-term challenges of managing diabetes effectively (McMahon et al., 2007; Polonsky et al., 2023; Yardley & Sigal, 2015).

In recent years, machine learning (ML) has emerged as a transformative tool in healthcare, offering new avenues for predictive analytics in diabetes care (Makroum et al., 2022). The predictive power of ML holds significant potential for predicting hypoglycaemia during exercise. By providing individuals with type 1 diabetes with information regarding their risk of hypoglycaemia before they begin exercising, they can take preemptive measures to prevent hypoglycaemia, increasing their confidence and safety during exercise. Research to date shows promise, with three existing studies effectively predicting hypoglycaemia during exercise (Bergford et al., 2023; Mosquera-Lopez et al., 2023; Reddy et al., 2019). However, these predictive models are often complex and rely on specialised data, making them difficult to implement outside research studies and are rarely used by patients.

Addressing these challenges, we aimed to implement an approach that not only provides accurate prediction of hypoglycaemia during exercise, but also prioritises model simplicity and usability. By utilising a comprehensive dataset that integrates patient data from multiple cohorts, our goal was to develop and validate a model that removes the complexities of previous ML-based hypoglycaemia prediction models, creating a user-friendly tool for individuals with type 1 diabetes to predict their risk of hypoglycaemia during exercise. This represents a step towards improving the real-world usability of predictive models, where quick and clear risk assessment can directly support safe exercise among those with type 1 diabetes.

## Methods

An overview of the workflow is provided in electronic supplementary material (ESM) Fig. 1 along with more detailed methods regarding the ML model building. All data analysis and processing were conducted using Pandas (McKinney, 2010) and numPy (Harris et al., 2020) in Python 3.9. All code for this project is openly available in a GitHub repository (https://github.com/cafoala/glucoseGo). Development and validation of ML models were conducted in accordance with the TRIPOD framework (Collins et al., 2015).

**Figure 1.**
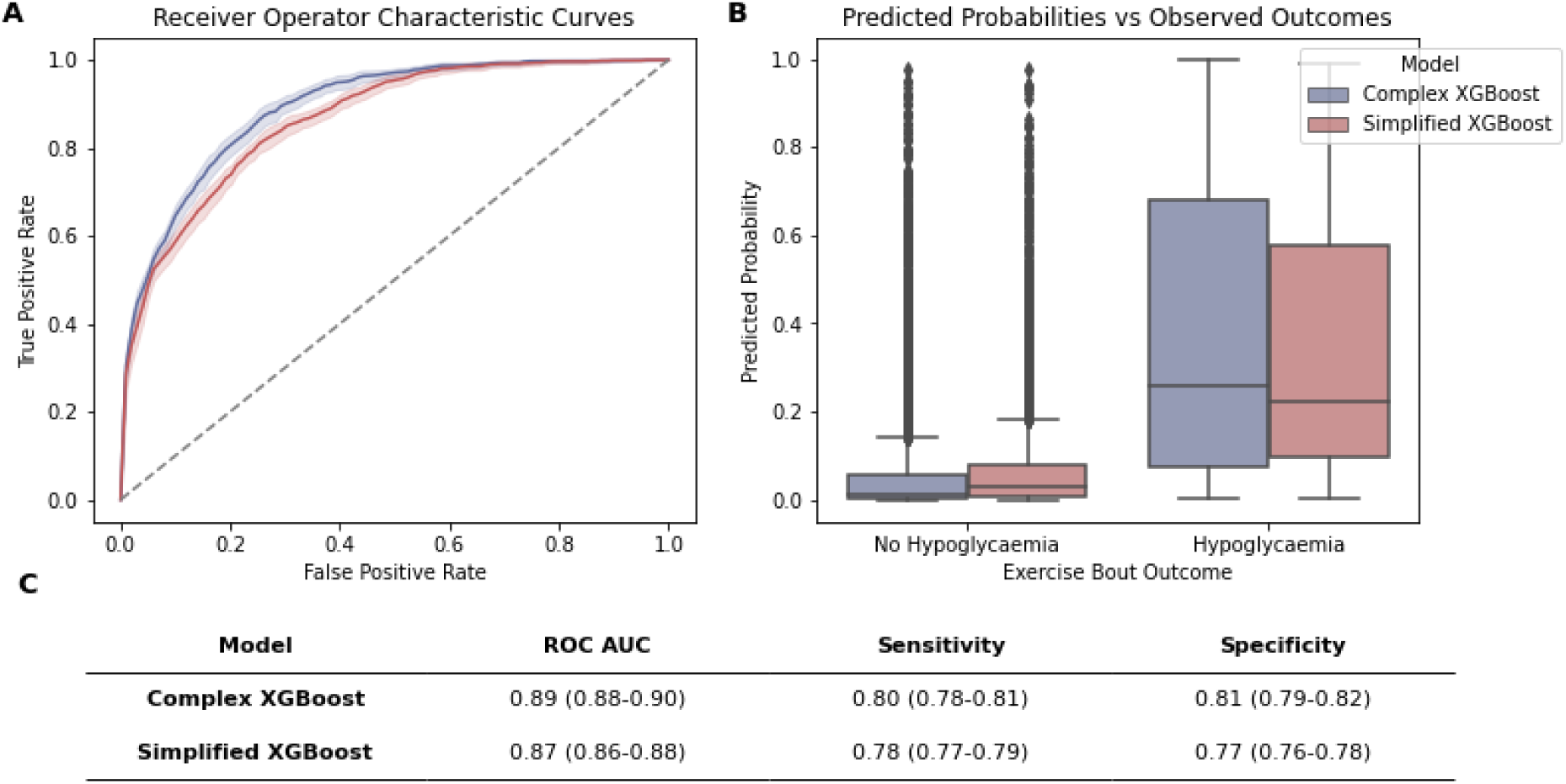
Accuracy measures for predicting hypoglycaemia during exercise. A: Receiver Operating Characteristic curve using all variables (blue) or two features (red), with the shaded area showing 95% confidence intervals (CIs). B: Distribution of predicted probability of hypoglycaemia for bouts that do and do not result in hypoglycaemia, divided into the two models. The dashed line represents the mean selected threshold for binary classification. C: Tabulated results for performance measures with 95% CIs. All CIs are derived from 10-fold cross-validation.

### Data source and participants

This study is a retrospective cohort analysis using four datasets. Two of these come from the EXercise in Type One Diabetes (EXTOD) group (https://www.extod.org/) and two from The Jaeb Center for Health Research (JCHR) (https://www.jaeb.org/). The EXTOD-101 (EXT-101) study followed 69 adults with type 1 diabetes training for a long-distance running race (Bunnewell et al., 2021). Glucose was recorded with a FreeStyle Libre continuous glucose monitor (CGM) and exercise bouts recorded in an exercise diary for six weeks prior to three weeks post-race. The EXTOD-Education (EXT-EDU) study (Narendran et al., 2020) assessed whether a type 1 diabetes educational programme improved glucose control around exercise. There were two weeks of data for 98 participants: one week at baseline and one week six months post-intervention. Glucose was recorded using a blinded Dexcom G6 CGM, and exercise bouts recorded with an exercise diary.

The JCHR datasets are from two large observational exercise studies, one in adults (Riddell et al., 2023) and one in youths (Riddell et al., 2024). Five hundred and sixty one adult participants in the Type 1 Diabetes EXercise Initiative (T1DEXI) study provided four weeks of free-living glucose, insulin and exercise data. Two hundred and fifty one young people (age 12-17 years) in the Type 1 Diabetes EXercise Initiative Pediatric (T1DEXIP) study provided 10 days of free-living glucose, insulin and exercise data. In both cohorts, participants wore an unblinded Dexcom G6 CGM and recorded exercise on a fitness tracker (Verily Study Watch for adults and Garmin Vivosmart for young people). The full details of the exercise protocols and data collection procedures have been detailed elsewhere (Riddell et al., 2023, Riddell et al., 2024).

### Inclusion criteria

Exercise bouts were excluded if: we could not determine the precise date/time that they occurred (N=71), they were duplicated or overlapping (N=831), they were outside of the range of 10 to 120 minutes in duration (N=2353), there was incomplete corresponding CGM data (<60% data sufficiency during bout and <40% data sufficiency for the hour prior) (N=2744), or the reported ‘exercise’ bouts were actually activities of daily living or occupational activities rather than volitional exercise as defined by Caspersen et al. (1985) (N=1167) (ESM Fig. 1).

### Predictor and target variables

Hypoglycaemia was the target variable and was determined using CGM data. It was defined as any readings below the threshold of level one hypoglycaemia (3.9 mmol/L or 70 mg/dL) during the exercise bout. This threshold was chosen because patients are advised to stop exercising and take corrective action when readings fall to this level.

We assessed the following predictor variables based on previous literature: age, sex, BMI (kg/m^2^), years since diagnosis, method of insulin administration (multiple daily injections [MDI], insulin pump, closed-loop system), HbA1c (mmol/mol), the exercise start time (morning, afternoon, or evening), exercise duration (minutes), the type of exercise (converted to predominantly aerobic, anaerobic, or mixed by domain expert), and the intensity (light, moderate, or vigorous).

For insulin administration, we extracted two variables: insulin on board per kg (IOB) and time since last insulin bolus dose (rounded to the nearest hour). IOB was calculated using a linear degradation algorithm with the duration of insulin action set to four hours.

From CGM data, the starting glucose was determined from the last reading before the start of exercise, and the glucose rate of change was computed using the difference between this reading and the glucose reading 15 minutes prior. The cut-offs for glucose rate of change were, Falling: <-0.05 mmol/L/min, Stable: -0.05 to 0.05 mmol/L/min, Rising: >0.05 mmol/L/min (E. M. Miller, 2020). For the one-hour CGM period prior to exercise, we used *Diametrics* (Russon et al., 2024) to extract standard metrics of glycemic control (e.g. time in normal range, average glucose, CV) (Danne et al., 2017) and *tsfresh* (Christ et al., 2018) to extract advanced statistical features from the CGM data. Ultimately, our dataset comprised 406 distinct variables and a total of 16,430 exercise bouts (ESM Table 1).

**Table 1.**
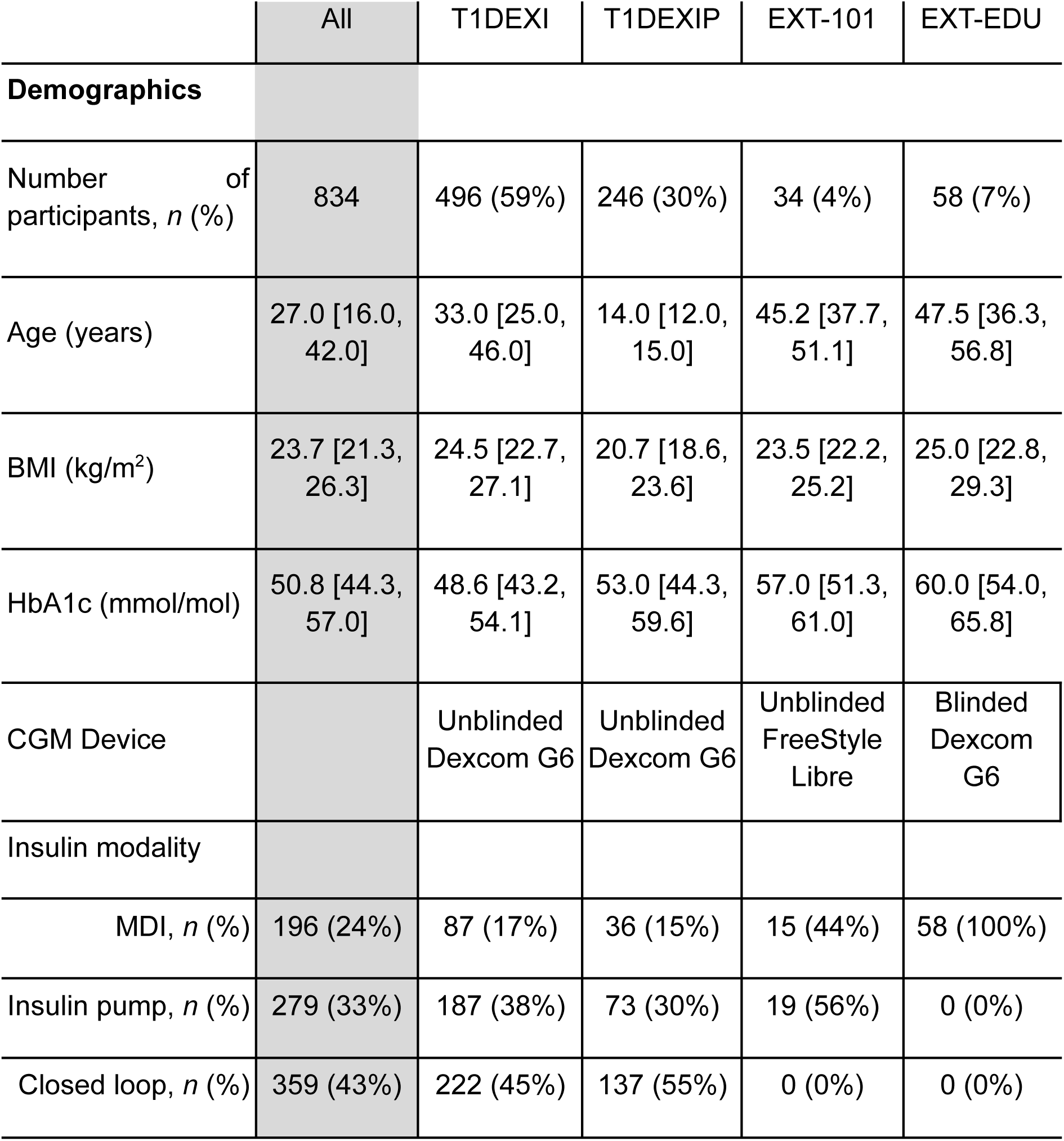
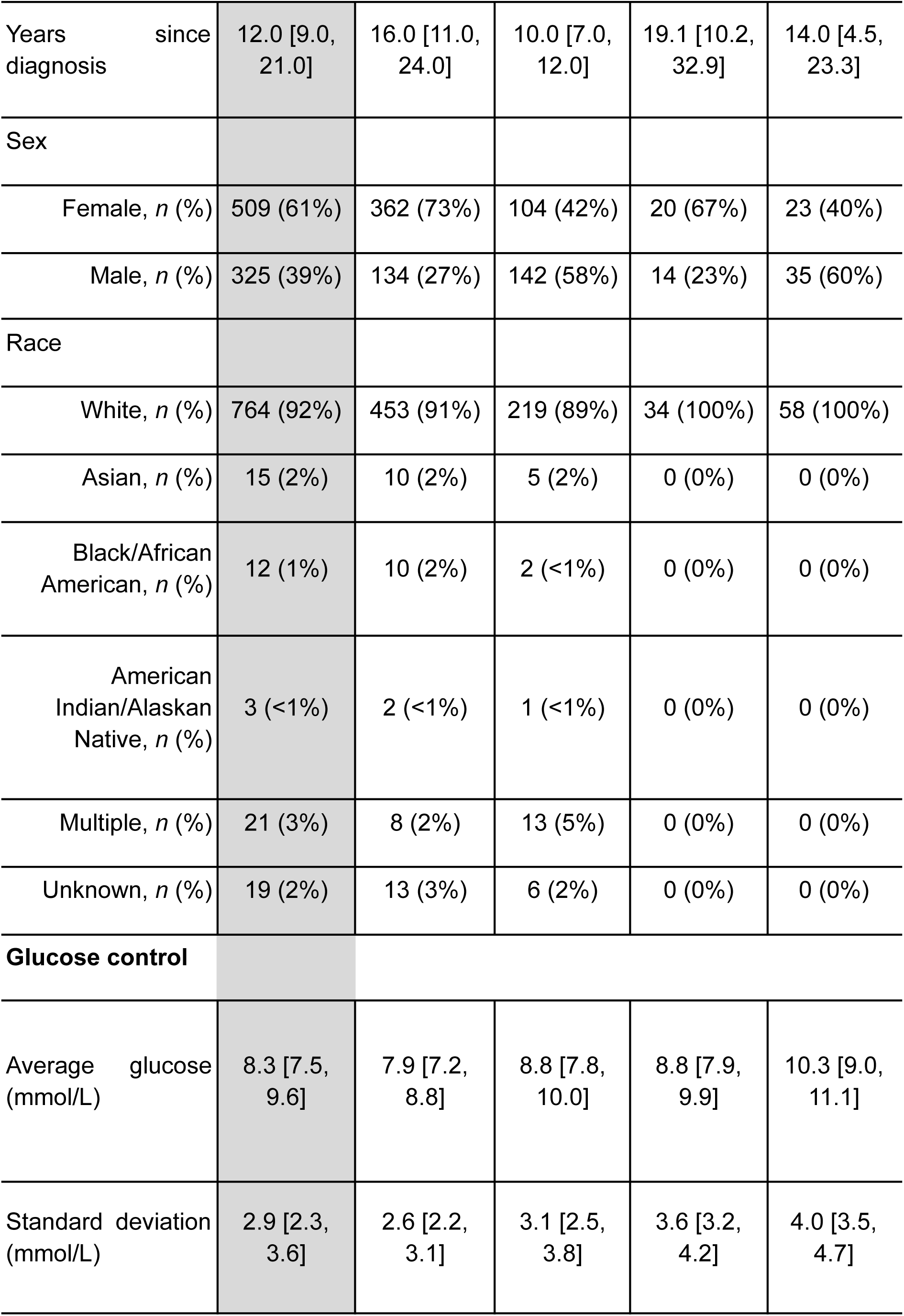
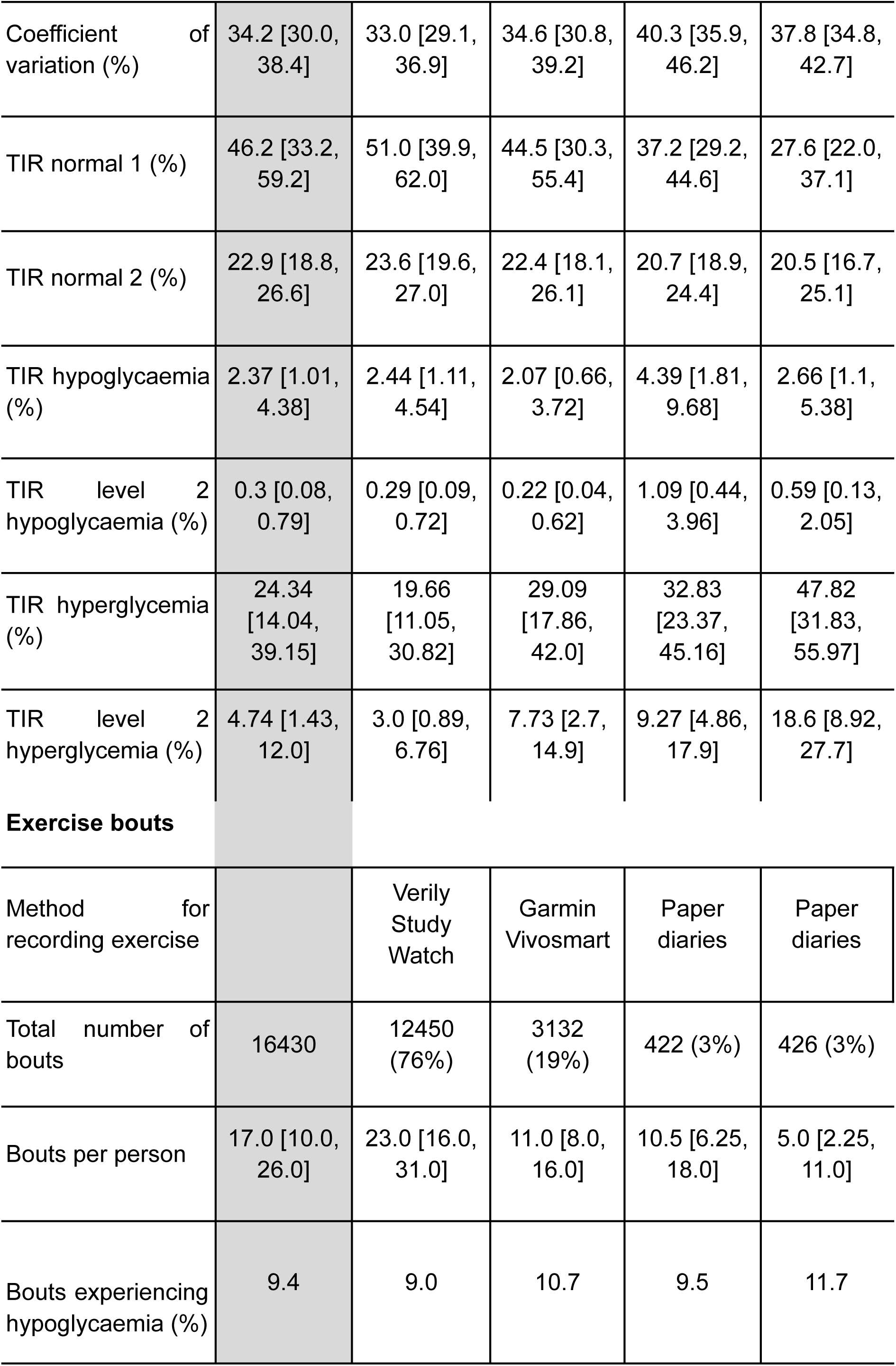
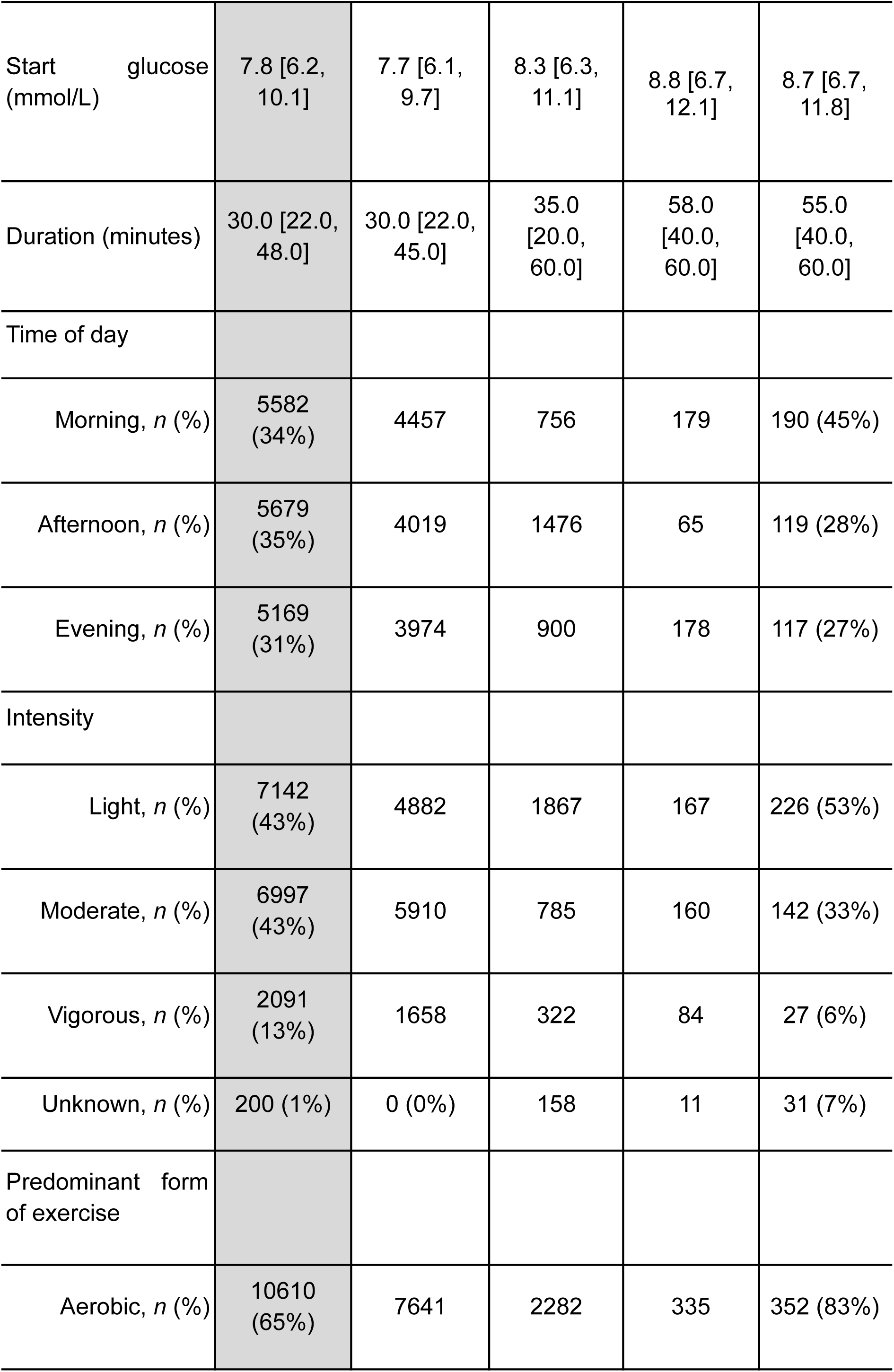
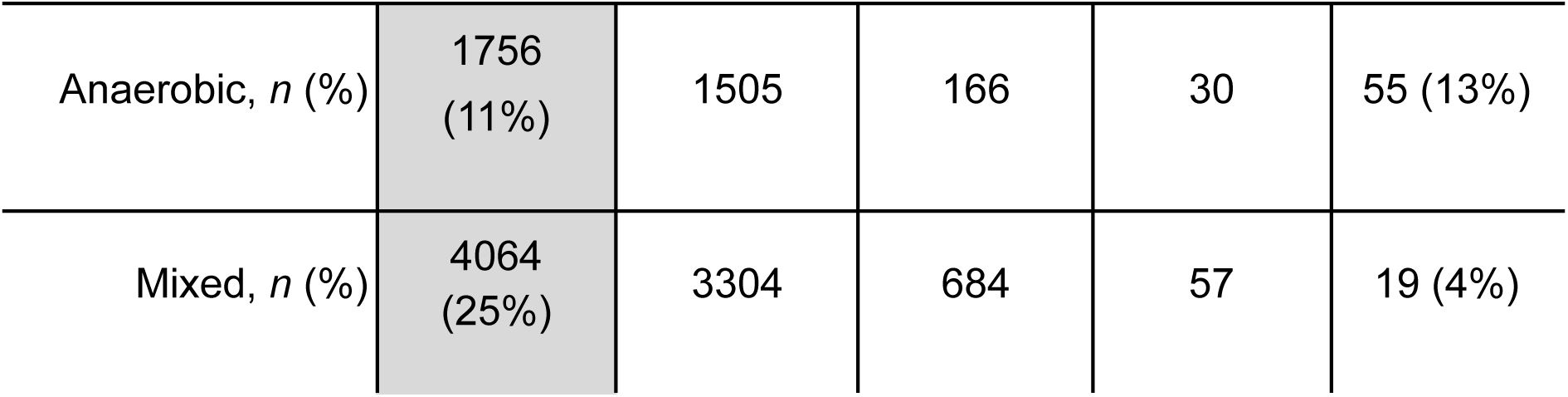
Characteristics of participants included in the study. Data are presented as median [IQR] unless otherwise specified. Percentages are rounded to the nearest whole number. Characteristics for the four study cohorts are detailed, alongside their aggregated characteristics (’All’).

|  | All | T1DEXI | T1DEXIP | EXT-101 | EXT-EDU |
| --- | --- | --- | --- | --- | --- |
| **Demographics** |  |  |  |  |  |
| Number of participants, *n* (%) | 834 | 496 (59%) | 246 (30%) | 34 (4%) | 58 (7%) |
| Age (years) | 27.0 [16.0, 42.0] | 33.0 [25.0, 46.0] | 14.0 [12.0, 15.0] | 45.2 [37.7, 51.1] | 47.5 [36.3, 56.8] |
| BMI (kg/m<sup>2</sup>) | 23.7 [21.3, 26.3] | 24.5 [22.7, 27.1] | 20.7 [18.6, 23.6] | 23.5 [22.2, 25.2] | 25.0 [22.8, 29.3] |
| HbA1c (mmol/mol) | 50.8 [44.3, 57.0] | 48.6 [43.2, 54.1] | 53.0 [44.3, 59.6] | 57.0 [51.3, 61.0] | 60.0 [54.0, 65.8] |
| CGM Device |  | Unblinded Dexcom G6 | Unblinded Dexcom G6 | Unblinded FreeStyle Libre | Blinded Dexcom G6 |
| Insulin modality |  |  |  |  |  |
| MDI, *n* (%) | 196 (24%) | 87 (17%) | 36 (15%) | 15 (44%) | 58 (100%) |
| Insulin pump, *n* (%) | 279 (33%) | 187 (38%) | 73 (30%) | 19 (56%) | 0 (0%) |
| Closed loop, *n* (%) | 359 (43%) | 222 (45%) | 137 (55%) | 0 (0%) | 0 (0%) |
| Years since diagnosis | 12.0 [9.0, 21.0] | 16.0 [11.0, 24.0] | 10.0 [7.0, 12.0] | 19.1 [10.2, 32.9] | 14.0 [4.5, 23.3] |
| Sex |  |  |  |  |  |
| Female, *n* (%) | 509 (61%) | 362 (73%) | 104 (42%) | 20 (67%) | 23 (40%) |
| Male, *n* (%) | 325 (39%) | 134 (27%) | 142 (58%) | 14 (23%) | 35 (60%) |
| Race |  |  |  |  |  |
| White, *n* (%) | 764 (92%) | 453 (91%) | 219 (89%) | 34 (100%) | 58 (100%) |
| Asian, *n* (%) | 15 (2%) | 10 (2%) | 5 (2%) | 0 (0%) | 0 (0%) |
| Black/African American, *n* (%) | 12 (1%) | 10 (2%) | 2 (<1%) | 0 (0%) | 0 (0%) |
| American Indian/Alaskan Native, *n* (%) | 3 (<1%) | 2 (<1%) | 1 (<1%) | 0 (0%) | 0 (0%) |
| Multiple, *n* (%) | 21 (3%) | 8 (2%) | 13 (5%) | 0 (0%) | 0 (0%) |
| Unknown, *n* (%) | 19 (2%) | 13 (3%) | 6 (2%) | 0 (0%) | 0 (0%) |
| **Glucose control** |  |  |  |  |  |
| Average glucose (mmol/L) | 8.3 [7.5, 9.6] | 7.9 [7.2, 8.8] | 8.8 [7.8, 10.0] | 8.8 [7.9, 9.9] | 10.3 [9.0, 11.1] |
| Standard deviation (mmol/L) | 2.9 [2.3, 3.6] | 2.6 [2.2, 3.1] | 3.1 [2.5, 3.8] | 3.6 [3.2, 4.2] | 4.0 [3.5, 4.7] |
| Coefficient of variation (%) | 34.2 [30.0, 38.4] | 33.0 [29.1, 36.9] | 34.6 [30.8, 39.2] | 40.3 [35.9, 46.2] | 37.8 [34.8, 42.7] |
| TIR normal 1 (%) | 46.2 [33.2, 59.2] | 51.0 [39.9, 62.0] | 44.5 [30.3, 55.4] | 37.2 [29.2, 44.6] | 27.6 [22.0, 37.1] |
| TIR normal 2 (%) | 22.9 [18.8, 26.6] | 23.6 [19.6, 27.0] | 22.4 [18.1, 26.1] | 20.7 [18.9, 24.4] | 20.5 [16.7, 25.1] |
| TIR hypoglycaemia (%) | 2.37 [1.01, 4.38] | 2.44 [1.11, 4.54] | 2.07 [0.66, 3.72] | 4.39 [1.81, 9.68] | 2.66 [1.1, 5.38] |
| TIR level 2 hypoglycaemia (%) | 0.3 [0.08, 0.79] | 0.29 [0.09, 0.72] | 0.22 [0.04, 0.62] | 1.09 [0.44, 3.96] | 0.59 [0.13, 2.05] |
| TIR hyperglycemia (%) | 24.34 [14.04, 39.15] | 19.66 [11.05, 30.82] | 29.09 [17.86, 42.0] | 32.83 [23.37, 45.16] | 47.82 [31.83, 55.97] |
| TIR level 2 hyperglycemia (%) | 4.74 [1.43, 12.0] | 3.0 [0.89, 6.76] | 7.73 [2.7, 14.9] | 9.27 [4.86, 17.9] | 18.6 [8.92, 27.7] |
| **Exercise bouts** |  |  |  |  |  |
| Method for recording exercise |  | Verily Study Watch | Garmin Vivosmart | Paper diaries | Paper diaries |
| Total number of bouts | 16430 | 12450 (76%) | 3132 (19%) | 422 (3%) | 426 (3%) |
| Bouts per person | 17.0 [10.0, 26.0] | 23.0 [16.0, 31.0] | 11.0 [8.0, 16.0] | 10.5 [6.25, 18.0] | 5.0 [2.25, 11.0] |
| Bouts experiencing hypoglycaemia (%) | 9.4 | 9.0 | 10.7 | 9.5 | 11.7 |
| Start glucose<br>(mmol/L) | 7.8 [6.2,<br>10.1] | 7.7 [6.1,<br>9.7] | 8.3 [6.3,<br>11.1] | 8.8 [6.7,<br>12.1] | 8.7 [6.7,<br>11.8] |
| Duration (minutes) | 30.0 [22.0,<br>48.0] | 30.0 [22.0,<br>45.0] | 35.0 [20.0,<br>60.0] | 58.0 [40.0,<br>60.0] | 55.0 [40.0,<br>60.0] |
| Time of day |  |  |  |  |  |
| Morning, *n* (%) | 5582<br>(34%) | 4457 | 756 | 179 | 190 (45%) |
| Afternoon, *n* (%) | 5679<br>(35%) | 4019 | 1476 | 65 | 119 (28%) |
| Evening, *n* (%) | 5169<br>(31%) | 3974 | 900 | 178 | 117 (27%) |
| Intensity |  |  |  |  |  |
| Light, *n* (%) | 7142<br>(43%) | 4882 | 1867 | 167 | 226 (53%) |
| Moderate, *n* (%) | 6997<br>(43%) | 5910 | 785 | 160 | 142 (33%) |
| Vigorous, *n* (%) | 2091<br>(13%) | 1658 | 322 | 84 | 27 (6%) |
| Unknown, *n* (%) | 200 (1%) | 0 (0%) | 158 | 11 | 31 (7%) |
| Predominant form<br>of exercise |  |  |  |  |  |
| Aerobic, *n* (%) | 10610<br>(65%) | 7641 | 2282 | 335 | 352 (83%) |
| Anaerobic, *n* (%) | 1756<br>(11%) | 1505 | 166 | 30 | 55 (13%) |
| Mixed, *n* (%) | 4064<br>(25%) | 3304 | 684 | 57 | 19 (4%) |

### Model implementation and evaluation

We used XGBoost (eXtreme Gradient Boosting algorithm) as the ML algorithm for this analysis. XGBoost is a decision tree-based ensemble learning method known for its accuracy, generalisability, and ability to capture nonlinearity and interactions between features (Chen & Guestrin, 2016).

The T1DEXI dataset (N=12450) was used to build the models using stratified *k*-fold cross validation (*k*=10) (ESM Fig. 2). To account for the repeated measures structure in the dataset - where each participant had multiple exercise events recorded - participants, rather than individual exercise events, were grouped within each fold, preserving within-participant correlations and preventing data leakage between folds. The models were then optimised using Bayesian hyperparameter tuning (Akiba et al., 2019; Bergstra et al., 2015).

**Figure 2.**
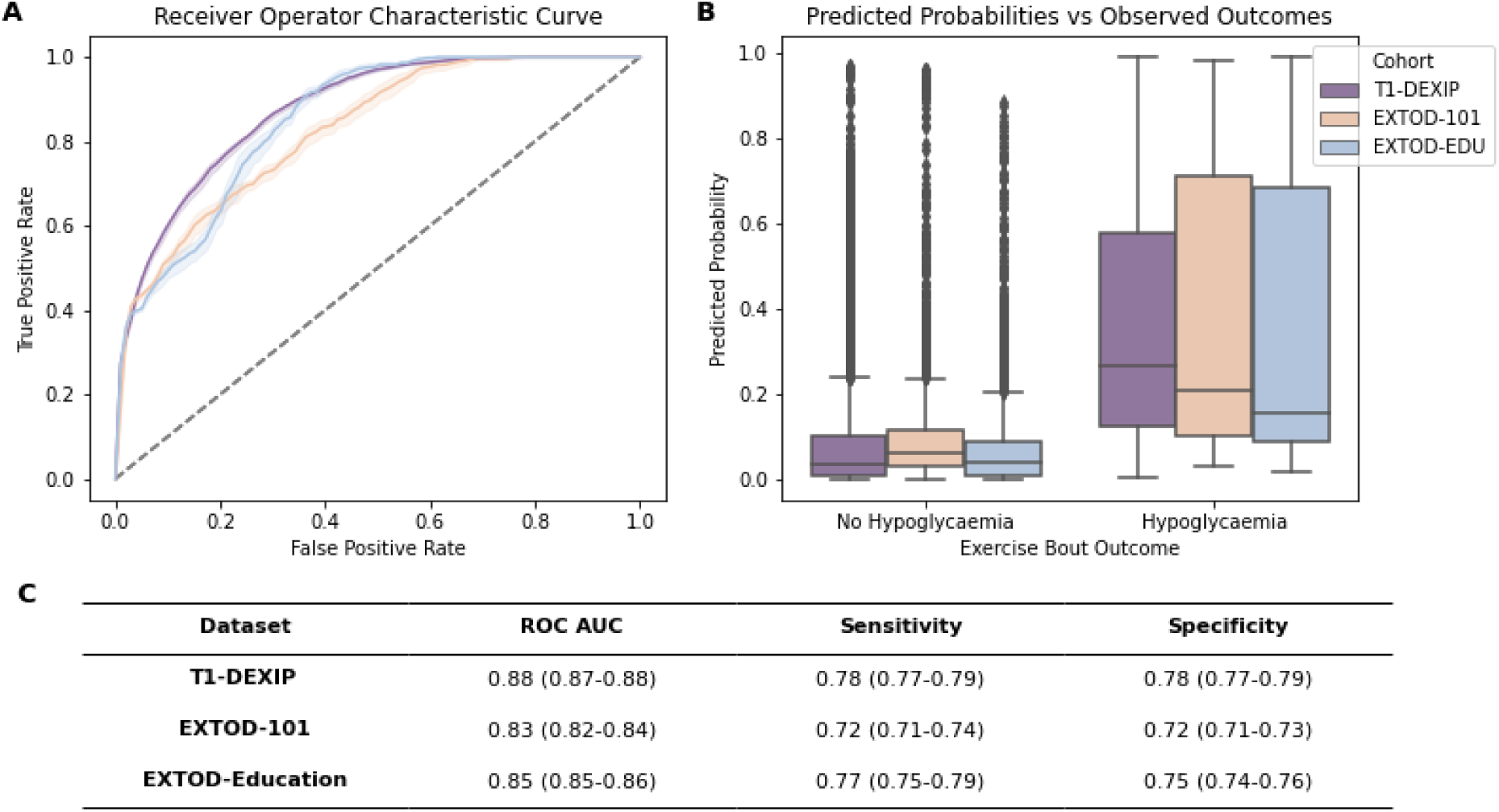
Validation of simplified model across independent cohorts. A: Receiver Operating Characteristic (ROC) curves for the simplified model in three independent cohorts: T1DEXIP (purple), EXTOD-101 (orange), and EXTOD-Education (blue), with shaded areas indicating 95% confidence intervals (CIs). B: Boxplots showing the distribution of predicted probabilities for bouts with and without hypoglycaemia across the three cohorts. The separation of the two distributions demonstrates the model’s discriminative performance. C: Tabulated results for performance measures with 95% CIs. All CIs are derived from 10-fold cross-validation.

The performance of the models were primarily assessed using the area under the receiver operating characteristics curve (ROC AUC) and calibration curve. In addition, we used thresholding to calculate sensitivity and specificity.

### Creating a simplified model

We identified 13 variables that are readily available to people with type 1 diabetes as they are about to begin exercise: type of exercise (aerobic, anaerobic, mixed), duration of exercise, intensity, time of day (morning (5AM-12PM), afternoon (12PM-5PM), and evening (5PM-5AM)), starting glucose, rate of change arrows (Falling: <-0.05mmol/L/min, Stable: -0.05-0.05mmol/L/min, Rising: >0.05mmol/L/min (E. M. Miller, 2020)), time since last insulin bolus (<1.5hrs ago, 1.5-3.5 hours ago, >3.5 hours ago), age, method of insulin administration, years since diagnosis, sex, BMI, and HbA1c. Using forward feature selection with XGBoost (Pedregosa et al., 2011), we iteratively assessed each feature’s contribution to model performance based on ROC AUC. To prioritise simplicity and usability as a visual tool, only the variables that we deemed to provide meaningful performance improvements without adding unnecessary complexity were selected. The model was then trained with only these selected variables, following the previously described methods.

### Assessing transferability to other populations

To test the transferability of the simplified model across different populations, we validated it on the three other cohorts: T1DEXIP (N=3132), EXTOD 101 (N=422), and EXTOD EDU (N=426). For each cohort, the simplified model was applied, and the mean prediction and 95% confidence intervals across folds was calculated to obtain a robust performance measure for each dataset.

The performance of the model on these separate cohorts was again evaluated using ROC AUC curves and sensitivity and specificity.

### Model transparency

To enhance the interpretability of XGBoost’s “black box” decisions, we utilised Shapley Additive Explanations (SHAP) values. This method assigns importance values to variables for individual predictions, providing a clearer understanding of their impact on the model’s outcomes (Lundberg & Lee, 2017).

We also conducted a subgroup analysis, calculating observed and predicted hypoglycaemia rates and ROC AUC, and visualised the results with forestPlot (Shen, 2023). The variables we assessed were starting glucose, duration of exercise, starting rate of change, IOB, age, sex, insulin administration, insulin modality, type of exercise, intensity of exercise, years since diagnosis, HbA1c, and CV. Race was also examined but we were unable to perform subgroup analysis due to the limited sample size across many of the racial subgroups.

## Results

### Participant characteristics

The characteristics of the participants are shown in Table 1. In total there were 834 participants, of which 61% were female. The median age of participants was 27 years old and duration of diabetes 12 years. Closed-loop systems were the most common insulin modality and overall glycemia was close to the recommended target of 48 mmol/mol for HbA1c and 70% for time in range (3.9-10 mmol/L), with median HbA1c being 50.8 mmol/mol and time in range being 69%.

There were 16430 recorded exercise bouts overall, with each person recording a median of 17 bouts of exercise. The median duration of an exercise bout was 30 minutes, and most bouts were of light intensity (43%) and primarily aerobic (65%). Across the entire dataset, 9.4% of exercise bouts resulted in hypoglycaemia. This was consistent across all studies. There were some differences between the cohorts, with EXTOD participants being, on average, older, majority on MDI, have higher HbA1c, and less time in range.

### Model Development: Forward Variable Selection and Simplified XGBoost Model

The results of forward feature selection are shown in Table 2. Starting glucose was identified as the most predictive variable, achieving an independent ROC AUC of 0.78. Adding exercise duration further increased the ROC AUC by 0.048, and the third variable, the “falling” rate of change arrow, improved it by an additional 0.023, up to 0.851.

**Table 2.** Variable Selection for XGBoost Model. This table presents the twelve variables identified as readily available to patients, ordered by importance, as determined through forward variable selection with an XGBoost model. The respective ROC AUC scores illustrate the impact on performance when each variable is added to the model.

| **Variables Included** | **ROC AUC Score** | **Change in ROC AUC score** |
| --- | --- | --- |
| Starting glucose (mmol/L) | 0.780 | 0.078 |
| + Duration (minutes) | 0.828 | 0.048 |
| + Rate of Change (Falling) | 0.851 | 0.023 |
| + Time since last insulin (>3.5 hrs) | 0.862 | 0.011 |
| + Time since last insulin (<1.5 hrs) | 0.865 | 0.003 |
| + Type of exercise (aerobic) | 0.867 | 0.002 |
| + Time of day (morning) | 0.871 | 0.004 |
| + Sex | 0.873 | 0.002 |
| + Time of day (evening) | 0.871 | -0.002 |
| Rate of Change (Stable) | 0.871 | 0 |
| + Type of exercise (mixed) | 0.871 | 0 |
| + Insulin modality (closed loop) | 0.869 | -0.002 |
| + Insulin modality (Pump) | 0.868 | -0.001 |
| + Intensity | 0.865 | -0.003 |
| + Age (years) | 0.853 | -0.012 |
| + Years since diagnosis | 0.846 | -0.007 |
| + BMI (kg/m<sup>2</sup>) | 0.843 | -0.003 |
| + HbA1c (mmol/mol) | 0.838 | -0.005 |

Subsequent variables contributed only marginal improvements (ROC AUC gains of <0.02). For instance, including time since the last insulin dose (>3.5 hours) increased the ROC AUC score by 0.011. A final improvement to a ROC AUC of 0.871 was achieved by adding time since the last insulin dose (<1.5 hours), type of exercise, and time of day. Beyond this point, incorporating more variables yielded no further improvements or even reduced performance, emphasising the diminishing returns of increased model complexity.

Ultimately, three key variables - starting glucose, exercise duration, and rate of change (classified as either Falling or Stable/Rising) - were selected for building the simplified XGBoost model. These variables captured the model’s predictive power while keeping it straightforward enough to enable the creation of a practical visual tool.

### Model Performance: Complex vs Simplified Model

The XGBoost model built with all 406 variables (“complex model”) predicted hypoglycaemia during exercise with high performance. The model achieved an ROC AUC of 0.89 (95% CI: 0.88–0.90) (Fig. 1A, blue line), a sensitivity of 0.80 (95% CI: 0.78–0.81) and a specificity of 0.81 (95% CI 0.79–0.82) (Fig. 1C). The XGBoost model trained on only three features (“simplified model”) achieved an ROC AUC of 0.87 (95% CI: 0.86–0.88) (Fig. 1A, red line); a 0.04 decrease compared to the complex model. The sensitivity was 0.78 (95% CI: 0.77–0.79) and the specificity was 0.77 (95% CI: 0.76–0.78).

The boxplot (Fig. 1B) illustrates the distribution of predicted probabilities for hypoglycaemia across the two outcomes, “No Hypoglycaemia” and “Hypoglycaemia”. The Hypoglycaemia group shows a greater spread of predicted probabilities compared to the tightly clustered, low probabilities in the No Hypoglycaemia group. Predicted probabilities are consistently higher for Hypoglycaemia, with minimal overlap between interquartile ranges, indicating strong model discrimination. Both models appear to effectively distinguish high- and low-risk exercise bouts.

The calibration curves (ESM Fig. 3) show how accurately the probability estimates of each model reflect the actual observed risk of hypoglycaemia in the data. The complex model (ESM Fig. 3, blue line) is generally well-calibrated, particularly at lower risk levels. At higher probability levels, it begins to show slight deviations, tending to overestimate risk of hypoglycaemia. The simplified model (ESM Fig. 3, red line) demonstrates superior calibration, aligning closely with the ideal calibration line across the full range of predicted probabilities. This suggests that the simplified model provides highly reliable probability estimates that more accurately reflect the actual risk of hypoglycaemia, and offer more interpretable and reliable risk estimates for clinical decision-making.

**Figure 3.**
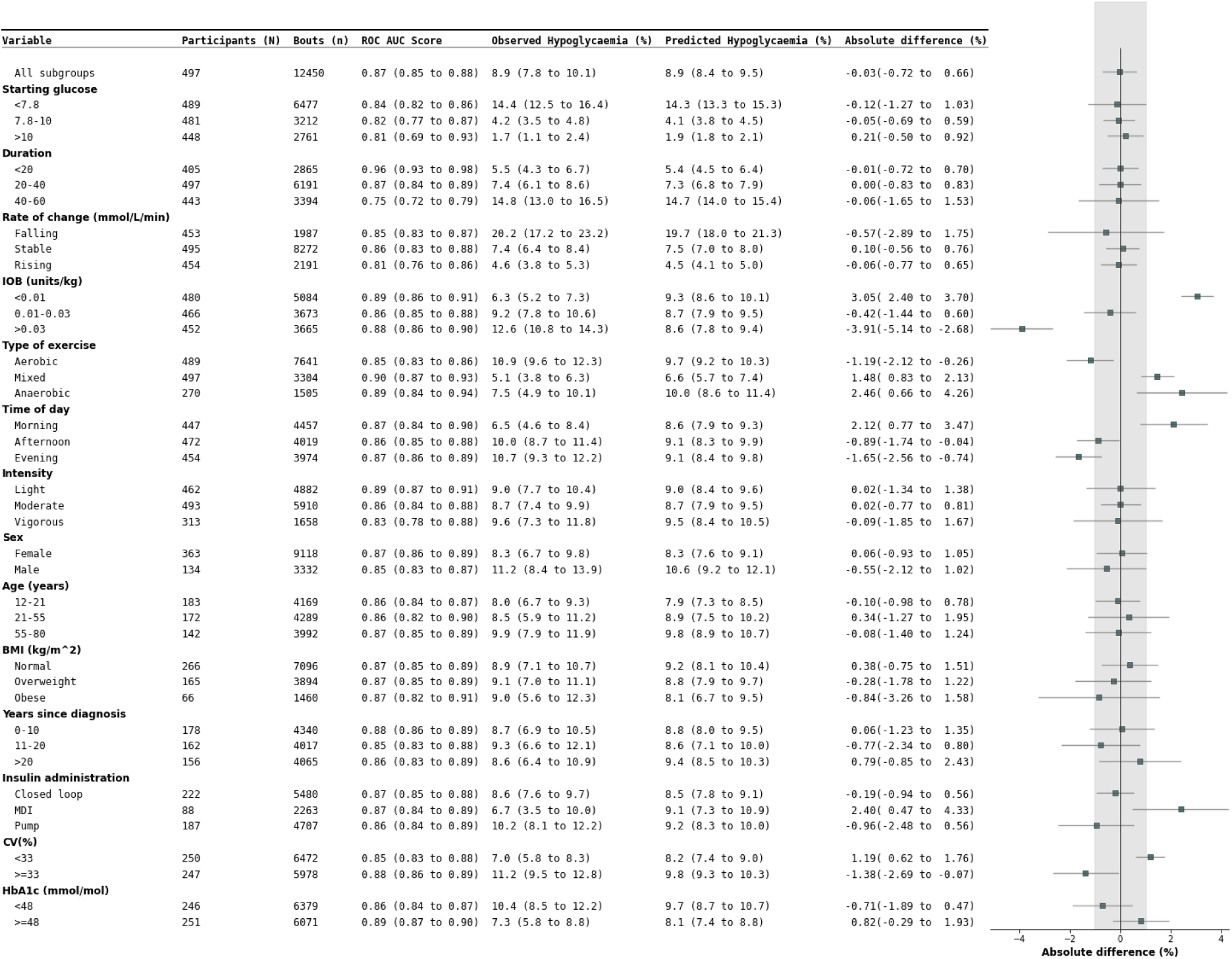
Subgroup Analysis of XGBoost Trained on Starting Glucose and Duration of Exercise. Each bar represents the ROC AUC score for the respective subgroup, with error bars indicating the 95% confidence intervals (CIs). Observed positive rate is the actual percentage of bouts in which hypoglycaemia occurs. Predicted positive rate is the mean predicted percentage of bouts in which hypoglycaemia occurs for each subgroup. ‘All subgroups’ is the overall ROC AUC score for the model, the grey dashed line and grey shaded area represents the mean ROC AUC and 95% CIs for the overall model, respectively.

SHAP dependence plots showing the relationships between clinically relevant features and hypoglycaemia risk are displayed in ESM Fig. 7 and 8. These plots replicate patterns observed in previous studies, such as the impact of starting glucose, exercise duration, type and intensity of exercise, while also highlighting the nonlinear contributions of many variables, validating XGBoost as a valid choice as model.

### Validation of Simplified Model Across Independent Cohorts

The simplified model was validated on three independent cohorts: T1DEXIP, EXTOD-101, and EXTOD-EDU, demonstrating excellent performance with mean ROC AUC scores ranging from 0.83 to 0.88 (Fig. 2A). The highest performance was observed in the T1DEXIP cohort (0.88 (95% CI: 0.87–0.88). The two EXTOD cohorts achieved the same mean ROC AUC: EXTOD-EDU cohort 0.85 (95% CI: 0.85–0.86) and EXTOD-101 cohort 0.83 (95% CI: 0.82–0.84).

Boxplots (Fig. 2B) show a clear separation in predicted probabilities between No Hypoglycaemia and Hypoglycaemia across all cohorts, underscoring the model’s strong discriminative ability. Calibration curves (ESM Fig. 4) further demonstrate that predicted probabilities closely align with observed outcomes. A slight overestimation of hypoglycaemia risk is noted in the EXTOD-101 cohort, particularly at higher probability levels, but overall, the model remains well-calibrated across all cohorts.

**Figure 4.**
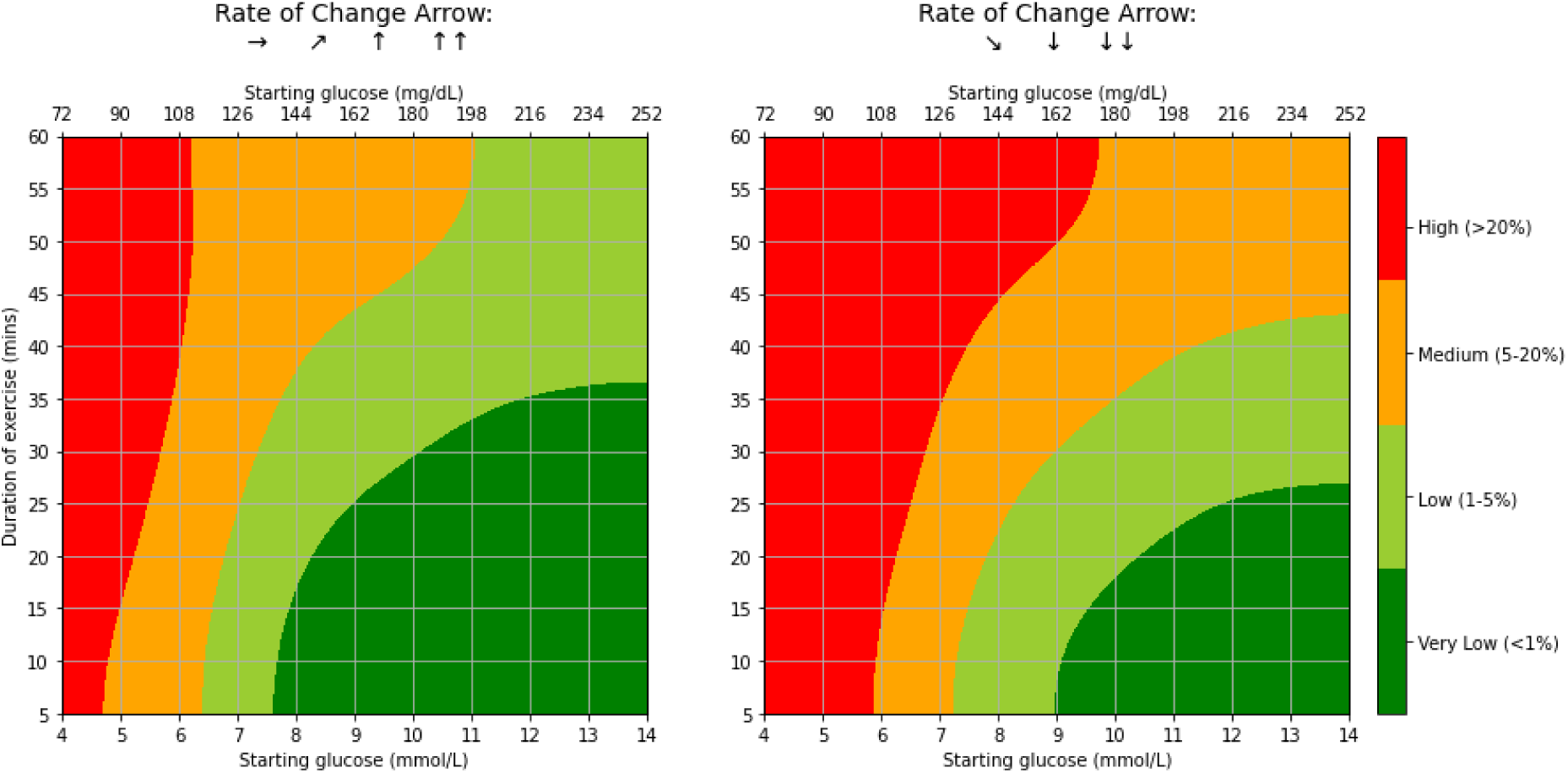
GlucoseGo Heatmap. Heatmap displaying the predicted risk of hypoglycaemia during exercise based on an XGBoost model trained on starting glucose levels and exercise duration. The colour-coded risk scores are represented as follows: dark green (<1% risk), green (1-5% risk), amber (5-20% risk), and red (>20% risk). The x-axis represents the starting glucose levels, while the y-axis represents the duration of the exercise.

### Subgroup analysis shows consistent predictive accuracy across participant subgroups

Subgroup analysis revealed notable variations in hypoglycaemia rates across subgroups (Fig. 3), with the most significant differences associated with key predictive factors such as starting glucose, glucose rate of change, exercise duration, and, to a lesser extent, IOB, exercise type, and time of day.

Despite these variations, the model’s predictions (Fig. 3, “Predicted Hypoglycaemia”) closely aligned with the observed hypoglycaemia rates (Fig. 3, “Observed Hypoglycaemia”), achieving an overall absolute difference of -0.03% (95% CI: -0.72%–0.66%). This indicates that the model’s predicted probabilities of hypoglycaemia closely correspond to the true likelihood of hypoglycaemia occurring in each subgroup.

This strong calibration was evident across most variables, including age, BMI, and sex. Minor discrepancies were observed for IOB, type of exercise and time of day. IOB had the greatest discrepancy, with the model underpredicting hypoglycaemia risk by a mean of 3.91% for high IOB levels and overpredicted risk by -3.05% for low IOB levels. Smaller discrepancies were seen for exercise type, with a slight underprediction during aerobic exercise (-1.19%) and overprediction during anaerobic (2.46%) and mixed activities (1.48%), and time of day, with underprediction for evening (-1.65%) and afternoon (-0.89%) exercise and overprediction for morning exercise (2.12%).

The model also demonstrated strong predictive performance across most subgroups, with ROC AUC scores closely aligning with the overall model’s ROC AUC and overlapping confidence intervals for most subgroups. The only exception was exercise duration, where model performance varied significantly. For durations under 30 minutes, the ROC AUC was 0.92 (95% CI: 0.89–0.94), indicating excellent predictive power. This dropped to 0.80 (95% CI: 0.78–0.82) for durations of 30–60 minutes and further to 0.69 (95% CI: 0.63–0.75) for durations over 60 minutes, reflecting a significant decline in predictive accuracy for longer bouts.

### Translation of Simple Model to a User-Friendly Heatmap

Using the simplified model, we were able to construct a simple heatmap as our patient-facing tool to illustrate predicted risk of hypoglycaemia according to starting glucose, exercise duration and rate of change arrows. Since performance decreases significantly after 60 minutes of exercise duration, we capped the heatmap here.

We developed a number of heatmaps in consultation with our diabetes Patient and Public Involvement (PPI) group. They felt that a four-colour traffic-light scheme with risk classifications of <1% for very low risk, 1-5% for low risk, 5-20% for medium risk, and >20% for high risk was the best way of representing this information. We named it the GlucoseGo heatmap, shown in Fig. 4.

The GlucoseGo heatmap was shown to be well calibrated. The observed positive rate for each traffic-light risk category falls comfortably within the bounds of the respective risk threshold (ESM Fig. 5). Those predicted to be green (<5% risk) had an observed hypoglycaemia rate of 1.8% (95%CI 1.5%-2.1%), amber (5-20% risk) of 11.5% (95%CI 10.7%-12.4%), and red (>20%) of 43.0% (95%CI 40.9%-45.1%). It was also shown to be well calibrated for the variables that showed the biggest variation in subgroup analysis. ESM Fig. 6 demonstrates that the model remained within acceptable bounds for these subgroups, with predictions falling within the boundaries of the heatmap categories.

## Discussion

In this study, we have effectively predicted hypoglycaemia during exercise using two XGBoost ML models on data from 834 participants from four different study cohorts, comprising 16,477 free-living exercise bouts. Starting glucose, exercise duration, and rate of change arrows (falling vs. stable/rising) emerged as the most important predictors of hypoglycaemia during exercise. Using just these three variables, a simplified XGBoost model achieved a mean ROC AUC of 0.87 (95% CI: 0.86–0.88). The complex model achieved an ROC AUC of 0.89 (95% CI: 0.88–0.90). The simplified model was translated into a traffic light risk-score heatmap. This provides people with type 1 diabetes an accurate, easy-to-use method for assessing their risk of hypoglycaemia during exercise, enabling them to exercise with more confidence.

The performance of our XGBoost models is consistent with previous studies predicting hypoglycaemia during exercise (Bergford et al., 2023; Mosquera-Lopez et al., 2023; Reddy et al., 2019). In line with earlier findings, our study confirms that glucose level at the start of exercise is the most crucial predictor of hypoglycaemia risk. Firstly, we found that both exercise duration and glucose rate of change are key predictors of hypoglycaemia risk - factors that are accessible to patients and can be easily assessed in real-world settings. Secondly, we were able to simplify our model to include only those variables that patients typically have on hand, without a substantial loss in predictive accuracy. This allowed us to create a practical, patient-facing tool that can be used in everyday life, moving beyond the complex or research-only models developed previously. Only one study created a simple, user-friendly tool (Reddy et al., 2019), but the applicability of this tool is limited by the small sample size and highly controlled settings.

The SHAP dependence plots reinforce relationships previously established in the scientific literature (Basu et al., 2014; Gomez et al., 2015; A. R. Jung et al., 2021; H. N. Jung et al., 2023; The Diabetes Research in Children Network (DirecNet) Study Group, 2006; Younk et al., 2011) and also clarify the model’s reliance on the three key variables. The overwhelming influence of starting glucose and exercise duration on the model’s predictions, compared to the relative unimportance of exercise type (aerobic, anaerobic, mixed) and intensity in the models was unexpected, given their recognised impact on hypoglycaemia risk during exercise. This discrepancy points to a potential divergence between clinical expectations and model-driven insights. Although it is possible that participants’ preemptive adjustments based on these factors might have influenced their lesser significance in the predictive models.

Based on the calibration curves, the simple XGBoost demonstrates better calibration than the complex XGBoost. This suggests that the simplified model may provide more reliable and interpretable probability estimates for clinical decision-making, as it avoids some of the potential overfitting seen in the more complex model.

The analysis of different patient subgroups demonstrated that the simplified model performed consistently well across a variety of groups, including those typically associated with unique glucose responses, such as young adults and individuals using closed-loop systems. This consistent efficacy across diverse patient groups is crucial in a clinical setting, ensuring that no particular group is at a disadvantage when using the model. Moreover, the finding that hypoglycaemia rates during exercise were similar among users of MDI, insulin pumps, and closed-loop systems highlights that, despite technological advances in diabetes management, there remains a significant need for straightforward, accessible tools to mitigate the risk of exercise-induced hypoglycaemia.

The simplified model demonstrated robust performance across diverse cohorts of participants with different ages (younger adults in T1-DEXIP compared to EXTOD), typical diabetes control (diabetes was less well controlled in EXTOD-101 and EXTOD-Education compared to T1-DEXIP), and cultural diversity (greater diversity in the British EXTOD cohorts compared to T1-DEXIP). This highlights the model’s potential utility across a range of demographic and clinical characteristics, suggesting its applicability to a broad population of people with type 1 diabetes. However, it is noteworthy that the model tends to slightly overestimate hypoglycaemia risk in the EXTOD-101 cohort. This may be attributable to the use of the FreeStyle Libre glucose monitoring system in this cohort, which records glucose readings every 15 minutes, compared to the 5-minute intervals used in other datasets. As we have reported previously the longer intervals between readings may reduce the system’s sensitivity to rapid glucose fluctuations, potentially affecting the identification of hypoglycemic events and influencing the calibration of the model’s risk predictions (Russon et al., 2025). Despite this, the simplified model maintained strong calibration and discriminative power, underscoring its potential as a simple, effective tool for predicting exercise-related hypoglycaemia risk in diverse clinical settings.

The GlucoseGo heatmap generated in this study, in collaboration with patients, holds considerable clinical promise, serving as a simple yet accurate tool for guiding exercise in individuals with type 1 diabetes. By distilling complex predictive factors into an easy-to-interpret format, the GlucoseGo heatmap not only empowers patients to engage in exercise with confidence, but also alleviates the mental burden often associated with managing the disease; commonly known as “diabetes burnout.” This is especially beneficial in sports or other settings where continuous monitoring may be impractical or intrusive.

The heatmap also aligns with clinical guidelines, reinforcing its reliability. It specifies a starting glucose threshold of 7 mmol/L (126 mg/dL) as the minimum for safely beginning exercise with a low risk of hypoglycaemia. This threshold corroborates the international position statement on the use and interpretation of CGMs around exercise (Moser et al., 2020), which also recommends this glucose level as a safe starting point for physical activity without the necessity of extra carbohydrate consumption.

It is however important to note that the heatmap’s development utilised data from free-living exercise bouts, where participants may have already adjusted their insulin or device settings to account for activity. Therefore, while the GlucoseGo heatmap is a valuable tool, it is meant to complement, not replace, standard pre-exercise precautions. After usual exercise preparations, individuals can refer to the heatmap to assess if further measures are necessary to avoid hypoglycaemia, enhancing their exercise safety and experience.

A major strength of this study is our dataset of over 16,000 free-living exercise bouts of varying type (aerobic, anaerobic and mixed), including participants with a wide age range (aged 12-80 year) who were in a variety of insulin delivery devices (MDI, insulin pump, closed-loop). In addition, rather than limiting the dataset by imposing strict criteria, we tried to use as much data as possible to ensure that we trained the ML algorithms on the most diverse dataset, as this has been shown to produce the most robust models (Belkin et al., 2019; Mandhala et al., 2022; Sharafutdinov et al., 2023).

This study has several limitations. The first is participant characteristics; the model has not been evaluated in children with diabetes under 12 years, it is largely a white ethnic cohort, and the diabetes cases have HbA1c and TIR close to recommended glycaemic targets. These factors could potentially limit the applicability of our findings to more ethnically or clinically diverse groups. Secondly, combining data from four different studies invariably results in inconsistencies in data collection methods. Specifically, the EXTOD studies relied on self-reported exercise intensity, the JAEB studies used device-derived metrics. Thirdly, the study employs a retrospective design, which makes it susceptible to biases stemming from pre-existing data, such as incomplete records or inconsistencies in data collection methods over time.

Further work is needed using a more extensive dataset encompassing a more ethnically and clinically diverse demographic. A key objective should be to assess this model’s performance within children with diabetes under the age of 12, where there could be significant potential benefits for caregivers and educators in physical education settings. It would also be helpful to do more PPI research to gain feedback to further refine the GlucoseGo heatmap. Finally, it would be ideal to validate the utility of the GlucoseGo heatmap through a clinical trial aimed at evaluating its efficacy in mitigating hypoglycaemia during exercise.

In summary, GlucoseGo represents a major advance in making hypoglycaemia risk prediction during exercise both accessible and practical for people living with type 1 diabetes. By combining cutting-edge machine learning with patient-centred design, we have delivered a simple, intuitive tool that can empower individuals to exercise more confidently and safely. This approach not only supports greater participation in physical activity, but also has the potential to reduce the burden of hypoglycaemia and improve overall quality of life. As GlucoseGo moves toward real-world implementation, it offers a promising pathway to transform diabetes self-management, encourage healthy behaviours, and drive further innovation at the intersection of digital health and diabetes care.

## Supporting information

ESM

## Data Availability

The T1DEXI and DEXIP datasets are available upon application from the Helmsley Charitable Trust (see https://helmsleytrust.org/request_for_proposal/improving-exercise-with-type-1-diabetes-moving-data-towards-solutions/ and follow data access instructions). The EXTOD datasets are available from the authors upon reasonable request and with permission of Prof Rob Andrews.

## Abbreviations

CGM: Continuous Glucose Monitor
EXTOD: EXercise in Type One Diabetes
EXT-101: EXTOD-101 study
EXT-EDU: EXTOD-Education study
IOB: Insulin on Board Units per kg
JCHR: Jaeb Center for Health Research
MDI: Multiple Daily Injections
PPI: Patient and Public Involvement
ROC AUC: Area Under the Receiver Operating Characteristics Curve
SHAP: SHapley Additive Explanations
T1DEXI: Type 1 Diabetes EXercise Initiative
T1DEXIP: Type 1 Diabetes EXercise Initiative Pediatric
XGBoost: eXtreme Gradient Boosting Algorithm

## Acknowledgements

Thank you to Angus Jones for critical evaluation of our findings.

This study was conducted with the support of the NIHR Exeter Biomedical Research Centre (BRC).

This publication is based on research using data from the Type 1 Diabetes EXercise Initiative (T1DEXI) and Type 1 Diabetes EXercise Initiative Pediatric (T1DEXIP) studies that has been made available through Vivli, Inc. Vivli has not contributed to or approved, and is not in any way responsible for, the contents of this publication.

During the course of preparing this work, the authors used ChatGPT and Copilot for the purpose of summarising research papers, text editing, and providing code snippets. Following the use of this tool/service, the authors formally reviewed the content for its accuracy and edited it as necessary. The authors take full responsibility for all the content of this publication.

## Contribution statement

All authors were involved in the conception, design, and conduct of the study, as well as the analysis and interpretation of the results. C.L.R. wrote the code and the first draft of the manuscript. M.J.A. served as the technical advisor for code and data analysis, R.C.A. acted as the clinical advisor, and R.M.P. provided expertise as the sports science advisor. All authors edited, reviewed, and approved the final version of the manuscript. C.L.R. is the guarantor of this work and, as such, had full access to all the data in the study and takes responsibility for the integrity of the data and the accuracy of the data analysis. The authors have no conflicts of interest.

