## Supplementary material for "GlucoseGo: A Simple, User-Friendly, Machine Learning-Derived Tool for Predicting Exercise-Related Hypoglycaemia Risk in Type 1 Diabetes": ESM

### 1. Methods Overview

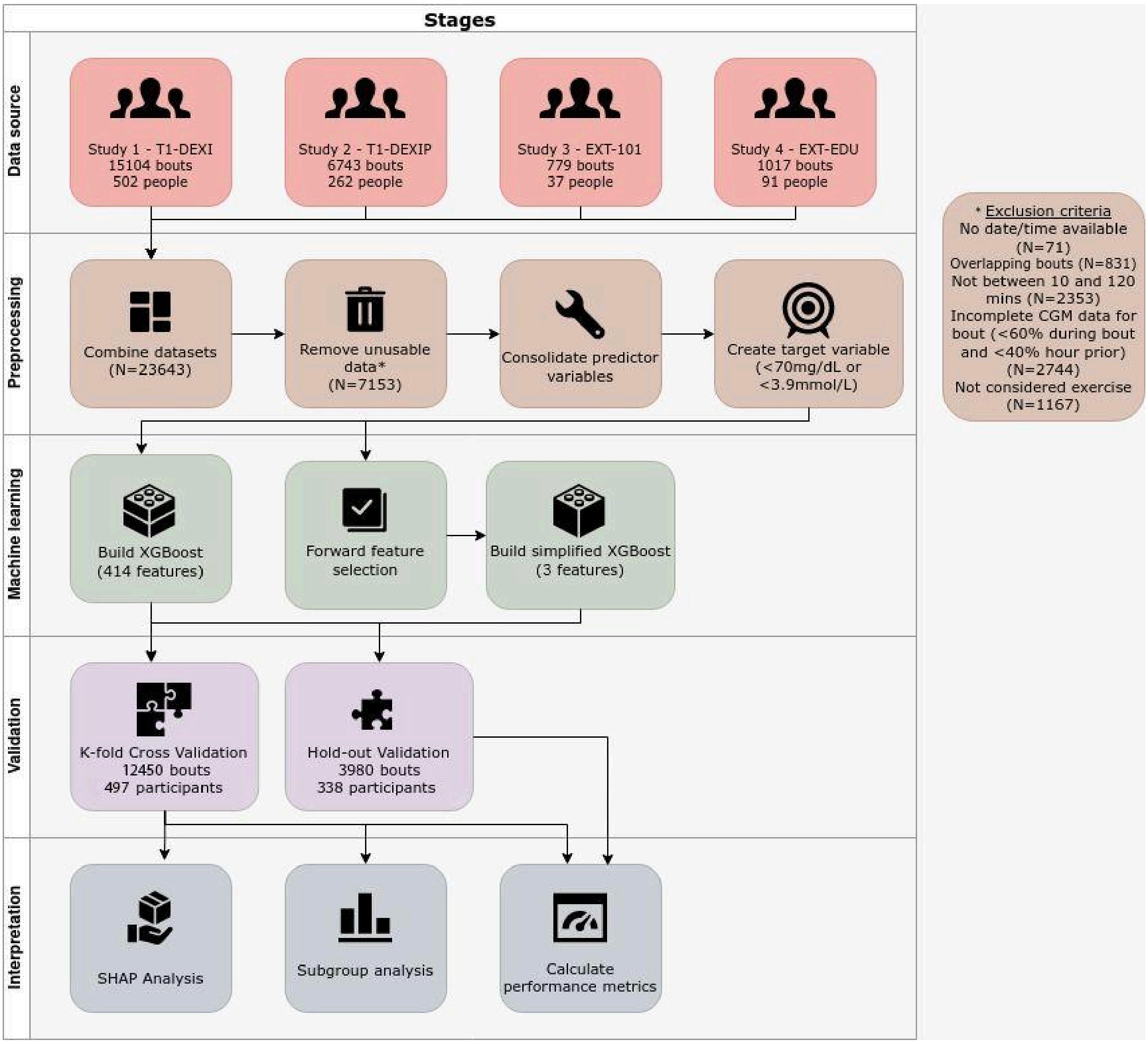


**Figure 1. Graphical Representation of Modelling Method, Including Data Exclusions.**

### 2. Data Dictionary

### A)

| Feature name | Description | Data type | Unique values | Missing values | Categories |
|---|---|---|---|---|---|
| intensity | The self-reported intensity of the exercise bout. 0=light, 1=moderate and 2=vigorous intensity. | Ordinal | 4 | 0 | 0, 1, 2 |

| | | | | | |
|---|---|---|---|---|---|
| day_of_week | The day of the week the exercise occurred. 0= Monday, 1=Tuesday, etc. | Categorical | 7 | 0 | 0, 1, 2, 3, 4, 5, 6, 7 |
| time_of_day | Time of day bout occurred. Morning 05:00-12:00, Afternoon 12:00-17:00, Evening 17:00-05:00 | Categorical | 3 | 0 | 'morning', 'afternoon', 'evening' , |
| form_of_exercise | The predominant type of the exercise | Categorical | 3 | 1 | 'aer', 'mix', 'ana' |
| sex | Sex | Categorical | 2 | 0 | 'male' , 'female' |
| insulin_modality | Type of insulin administration | Categorical | 3 | 11 | 'mdi' , 'pump' , 'closed_loop' |
| season | The season in which the exercise occurred | Categorical | 4 | 0 | 0, 1, 2, 3 |
| ID | Identifies participants using the study name and a unique number | Categorical | 834 | 0 | ‘dexip_100' … ‘dexi_988' |
| stratify | Variable created to stratify the data based on ID and target variable | Categorical | 1356 | 0 | ‘dexip_100_False' ... ‘dexi_988_True' |
| study | Which study the participant is in | Categorical | 4 | 0 | 'dexip', 'ext_101', 'ext_edu', 'dexi' |
| bout_id | The ID for individual bouts, ID plus timestamp of bout start | Categorical | 16577 | 0 | 'dexip_100_20210610130600' ... 'dexi_988_20213312083306' |
| y | Target variable – CGM dropped below 3.9mmol/L or 70mg/dL during exercise | Boolean | 2 | 0 | False, True |

**B)**

| Feature name | Description | Units | Missing values (N) | Min. value | Q1 | Q2 | Q3 | Max. value |
|---|---|---|---|---|---|---|---|---|
| duration | Duration of exercise bout | minutes | 0 | 10.0 | 22.0 | 30.0 | 48.0 | 120.0 |
| start_glc | The last glucose reading before exercising | mmol/L | 0 | 2.1 | 6.2 | 7.8 | 10.1 | 22.6 |
| start_roc | The rate of change before the last glucose reading and the reading >20mins before that reading | mmol/L/hour | 92 | -23.1 | -1.8 | 0.00 | 1.7 | 21.6 |

| | | | | | | | | |
|---|---|---|---|---|---|---|---|---|
| iob_kg | The insulin on board (IOB) per kg. Calculated using a linear degradation algorithm with duration of action set to 4 hours. | Units/kg | N/A | 0 | | | | |
| age | Participant's age | years | 27 | 12.00 | 21.00 | 30.00 | 46.00 | 80.16 |
| hba1c | HbA1c | mmol/mol | 136 | 29.0 | 43.2 | 48.6 | 55.2 | 140.4 |
| bmi | BMI | kg/m$^2$ | 39 | 13.7 | 21.6 | 23.7 | 26.3 | 48.5 |
| years_since_diagnosis | Years since diagnosis | years | 206 | 0.48 | 10.0 | 14.0 | 23.0 | 66.0 |

**C)**

| | Description | Units | Missing values (N) | Min. value | Q1 | Q2 | Q3 | Max. value |
|---|---|---|---|---|---|---|---|---|
| Data suff | The percentage of data present (total expected CGM readings minus missing readings) | % | 0 | 7.70 | 92.30 | 92.30 | 92.30 | 100.00 |
| Average glucose | The mean glucose of all the readings | (mmol/L) | 0 | 2.17 | 6.16 | 7.75 | 9.89 | 25.65 |
| eA1c | Estimated A1C | % | 0 | 2.99 | 5.51 | 6.50 | 7.85 | 17.76 |
| SD | Standard deviation | (mmol/L) | 11 | 0.00 | 0.34 | 0.59 | 1.00 | 5.23 |
| CV | Coefficient of variation | % | 11 | 0.00 | 4.27 | 7.43 | 12.82 | 72.26 |
| AUC | Area under the curve | mmol h/L | 15 | 0.31 | 2.76 | 3.76 | 5.52 | 20.42 |
| LBGI | Low blood glucose index | | 0 | 0.00 | 0.00 | 0.00 | 0.31 | 38.34 |
| HBGI | High blood glucose index | | 0 | 0.00 | 0.12 | 2.03 | 7.68 | 70.83 |
| MAGE | Mean amplitude of glycemic excursions | mmol/L | 28 | 0.05 | 0.83 | 1.50 | 2.61 | 14.05 |
| TIR normal | Percentage time in range 3.9-10mmol/L | % | 0 | 0.00 | 50.00 | 100.00 | 100.00 | 100.00 |
| TIR normal 1 | Percentage time in range 3.9-7.8mmol/L | % | 0 | 0.00 | 0.00 | 50.00 | 100.00 | 100.00 |
| TIR normal 2 | Percentage time in range 7.8-10mmol/L | % | 0 | 0.00 | 0.00 | 0.00 | 41.67 | 100.00 |
| TIR level 1 hypoglycemia | Percentage time in range 3.9-3.0mmol/L | % | 0 | 0.00 | 0.00 | 0.00 | 0.00 | 100.00 |
| TIR level 2 hypoglycemia | Percentage time in range <3.0mmol/L | % | 0 | 0.00 | 0.00 | 0.00 | 0.00 | 100.00 |
| TIR level 1 hyperglycemia | Percentage time in range 10.0-13.9mmol/L | % | 0 | 0.00 | 0.00 | 0.00 | 25.00 | 100.00 |
| TIR level 2 hyperglycemia | Percentage time in range >13.9mmol/L | % | 0 | 0.00 | 0.00 | 0.00 | 0.00 | 100.00 |

| | | | | | | | | |
|---|---|---|---|---|---|---|---|---|
| Total number hypoglycemic events | The number of times glucose drops below 3.9mmol/L for 15 mins or more | N | 0 | 0.00 | 0.00 | 0.00 | 0.00 | 1.00 |
| Number LV1 hypoglycemic events | The number of times glucose drops below 3.9mmol/L for 15 mins or more but not below 3.0mmol/L | N | 0 | 0.00 | 0.00 | 0.00 | 0.00 | 1.00 |
| Number LV2 hypoglycemic events | The number of times glucose drops below 3.0mmol/L for 15 mins or more | N | 0 | 0.00 | 0.00 | 0.00 | 0.00 | 1.00 |
| Number prolonged hypoglycemic events | The number of times glucose drops below 3.0mmol/L for 120 mins or more | N | 0 | 0.00 | 0.00 | 0.00 | 0.00 | 0.00 |
| Avg. length of hypoglycemic events | The mean length of all hypoglycemic episodes (anything below 3.9mmol/L) | minutes | 0 | 0.00 | 0.00 | 0.00 | 0.00 | 55.08 |
| Total time spent in hypoglycemic events | The combined length of all hypoglycemic episodes (anything below 3.9mmol/L) | minutes | 0 | 0.00 | 0.00 | 0.00 | 0.00 | 55.08 |
| Total number hyperglycemic events | The number of times glucose rises above 10.0mmol/L for 15 mins or more | minutes | 0 | 0.00 | 0.00 | 0.00 | 1.00 | 2.00 |
| Number LV1 hyperglycemic events | The number of times glucose rises above 10.0mmol/L for 15 mins or more but not above 13.9mmol/L | N | 0 | 0.00 | 0.00 | 0.00 | 0.00 | 2.00 |
| Number LV2 hyperglycemic events | The number of times glucose rises above 13.9mmol/L for 15 mins or more | N | 0 | 0.00 | 0.00 | 0.00 | 0.00 | 1.00 |
| Number prolonged hyperglycemic events | The number of times glucose rises above 13.9mmol/L for 120 mins or more | N | 0 | 0.00 | 0.00 | 0.00 | 0.00 | 0.00 |
| Avg. length of hyperglycemic events | The mean length of all hypoglycemic episodes (anything above 10.0mmol/L) | minutes | 0 | 0.00 | 0.00 | 0.00 | 24.98 | 59.93 |
| Total time spent in hyperglycemic events | The combined length of all hypoglycemic episodes (anything above 10mmol/L) | minutes | 0 | 0.00 | 0.00 | 0.00 | 24.98 | 59.93 |

**D:** Tsfresh Features Extracted from 1-Hour Pre-Exercise CGM Data

- Variance larger than standard deviation
- Has duplicate maximum

- Has duplicate minimum
- Has duplicate values
- Sum of values
- Absolute energy
- Mean absolute change
- Mean change
- Mean second derivative (central)
- Median
- Mean
- Length (number of readings)
- Standard deviation
- Coefficient of variation
- Variance
- Skewness
- Kurtosis
- Root mean square
- Absolute sum of changes
- Longest strike below mean
- Longest strike above mean
- Count above mean
- Count below mean
- Last location of maximum
- First location of maximum
- Last location of minimum
- First location of minimum
- Percentage of reoccurring values to all values
- Percentage of reoccurring datapoints to all datapoints
- Sum of reoccurring values
- Sum of reoccurring datapoints
- Ratio of value number to time series length
- Maximum
- Absolute maximum
- Minimum
- Benford correlation
- Time reversal asymmetry statistic (lags 1, 2, 3)
- C3 statistic (lags 1, 2, 3)
- Complexity-invariant distance (with and without normalization)
- Symmetry looking (various r values)
- Large standard deviation (various r values)
- Quantiles (10%, 20%, 30%, 40%, 60%, 70%, 80%, 90%)
- Autocorrelation (lags 0–9)
- Aggregated autocorrelation (mean, median, variance, max lag 40)
- Partial autocorrelation (lags 0–5)
- Number of CWT peaks (n=1, 5)
- Number of peaks (n=1, 3, 5, 10, 50)
- Binned entropy (10 bins)
- Index mass quantiles (various quantiles)
- Continuous wavelet transform (CWT) coefficients (various coefficients and widths)

- Welch's density coefficients (coeffs 2, 5)
- AR coefficients (coeff 10, k=10)
- Change quantiles (various quantiles and aggregation functions)
- Fast Fourier transform (FFT) coefficients (real, imaginary, absolute, angle, various coeffs)
- Aggregated FFT features (centroid, variance)
- Value count for specific values (0, 1, -1)
- Range count for specific max/min
- Approximate entropy (various m and r)
- Linear trend (p-value, r-value, intercept, slope, stderr)
- Aggregated linear trend (r-value, intercept, slope, stderr; chunk lengths 5/10; max, min, mean, variance)
- Augmented Dickey-Fuller test (test statistic, p-value, used lag)
- Number crossing mean (for various m)
- Energy ratio by chunks (num segments 10, segment focus 0–9)
- Ratio beyond r sigma (various r values)
- Count above a threshold (t=0)
- Count below a threshold (t=0)
- Lempel-Ziv complexity (various bin counts)
- Fourier entropy (various bin counts)
- Permutation entropy (dimension 3–7, tau=1)
- Mean of the n absolute maxima (n=7)

***Table 1. Data Dictionary Describing All Variables Used in Analysis.***
*A: Demographic, lab, and exercise data for categorical variables B: Demographic, lab, and exercise data for numerical variables. C: Metrics of glycemic control from CGM data one hour prior to exercise. D: List of time-series features extracted using tsfresh from the CGM trace in the hour preceding exercise.*

### 3. Missing data

To fill missing data, we used multivariate imputation using a *k*-nearest neighbour (*k*-nn) algorithm (*k*=5) (Pedregosa et al., 2011).

### 4. k-fold cross validation

The *k*-fold cross validation partitions data into 10 distinct subsets. For each iteration, a model is trained on 9 of these subsets and is validated on the remaining one, cycling through all subsets for comprehensive model validation (Wong & Yeh, 2020; Yadav &

Shukla, 2016). This results in 10 distinct models, with each data point in one, and only one, test set.

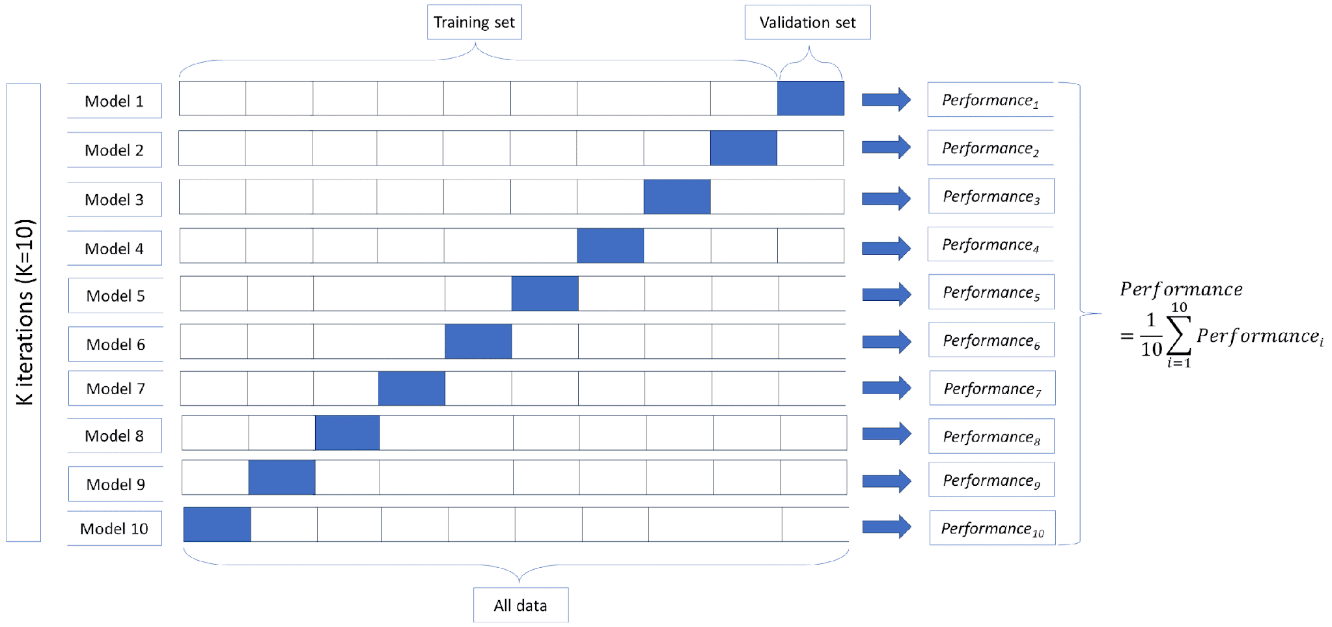


**Figure 2. *k*-fold Cross Validation.** Visual representation of *k*-fold cross validation (*k*=10).

### 5. Other Cohorts

For each cohort, the 10 *k*-fold simplified models were applied individually, and the mean prediction and 95% confidence intervals across folds was calculated to obtain a robust performance measure for each dataset.

### 6. Heatmap Construction

To construct the heatmap, we used the simplified model (10 *k*-folds) to produce a mean risk score for each grid point in the range of 4-14 mmol/L (72-252 mg/dL) and durations from 5-60 minutes. These predictions were smoothed using a Gaussian filter (Virtanen et al., 2020) followed by bicubic interpolation (Hunter, 2007).

### 7. Calibration of Models

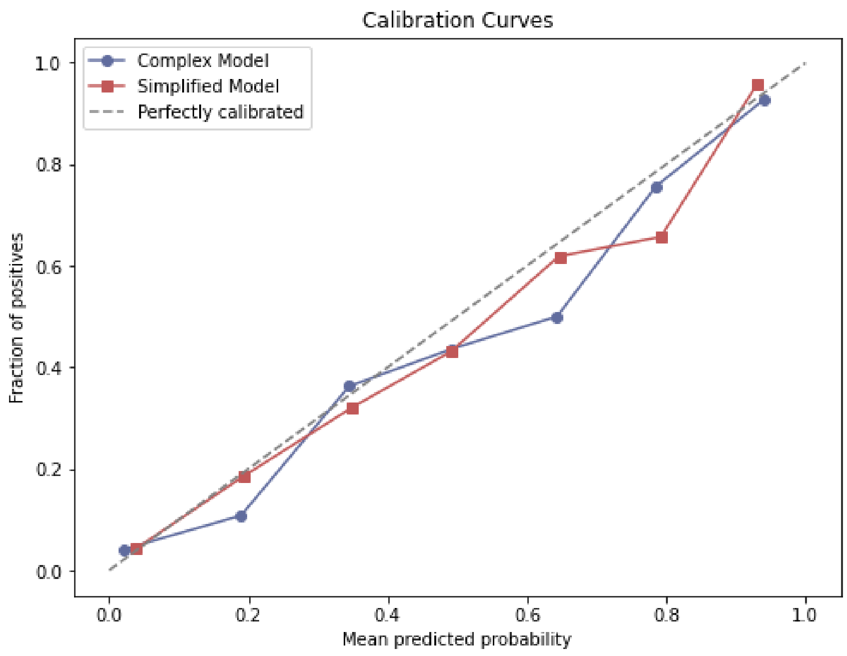


***Figure 3. Calibration curves for the complex and simplified models in the T1DEXI dataset.*** *The plot shows the calibration curves for the complex (blue line) and simplified (red line) XGBoost models using the T1DEXI dataset. The x-axis represents the predicted probability of hypoglycaemia during exercise, while the y-axis shows the observed proportion of exercise bouts that actually resulted in hypoglycaemia. The dashed diagonal line indicates perfect calibration, where predicted probabilities exactly match observed outcomes. Both models are generally well-calibrated, but the simplified model closely follows the ideal line across the full range of probabilities.*

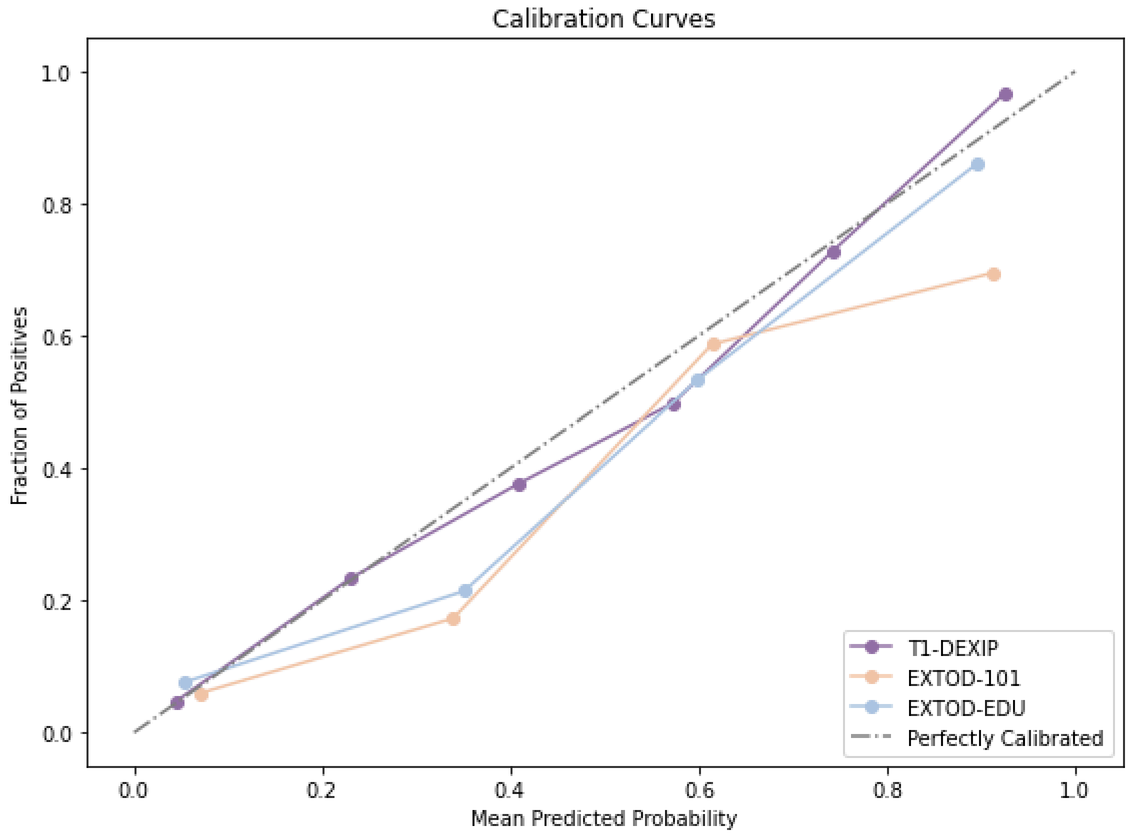


***Figure 4. Calibration curves for the simplified model in external validation cohorts.*** *The plot displays calibration curves for the simplified model applied to three independent validation cohorts: T1-DEXIP (purple line), EXTOD-101 (beige line), and EXTOD-EDU (blue line). The x-axis indicates the predicted probability of hypoglycaemia, and the y-axis represents the observed proportion of hypoglycaemia events in each probability bin. The dashed diagonal line corresponds to perfect calibration. The model remains well-calibrated across all cohorts, with a slight tendency to overestimate risk at higher probabilities in the EXTOD-101 cohort.*

**A**

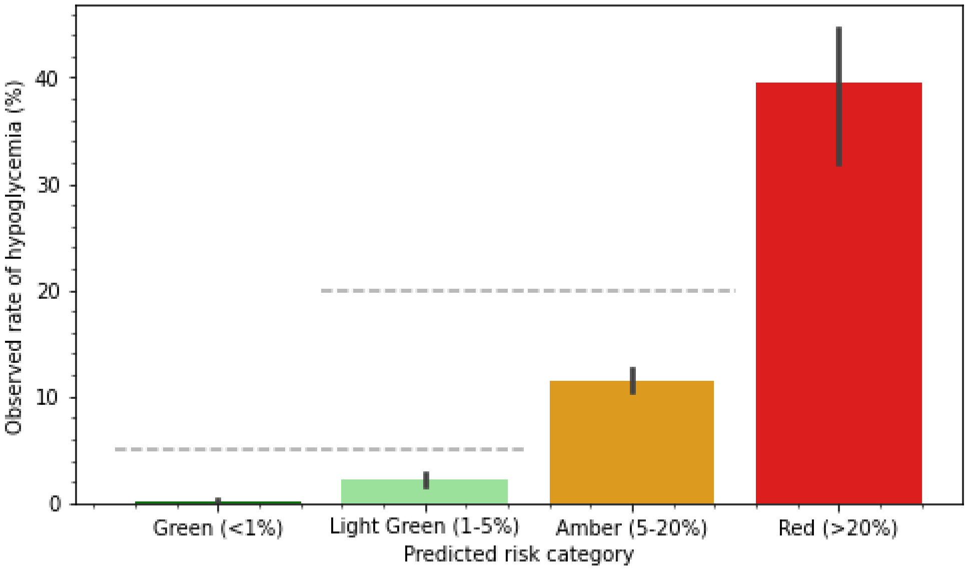


**B**

| **Predicted risk category** | **Observed rate of hypoglycemia (%)** |
|---|---|
| Green (<1%) | 0.2 (-0.0, 0.4) |
| Light Green (1-5%) | 2.2 (1.6, 2.7) |
| Amber (5-20%) | 11.5 (10.6, 12.4) |
| Red (>20%) | 39.5 (32.8, 46.3) |

***Figure 5. Calibration of heatmap results.*** *A: The bar chart shows the actual frequency of hypoglycaemia in the three heat-map risk zones of <1% (green), 1-5% (light green), 5-20%, (amber), and >20% (red). The grey dotted lines show the 5% and 20% thresholds. The spots represent the result for each of the 10 k-folds. B: The predicted and observed rate of hypoglycemia within the zones with the 95% confidence intervals.*

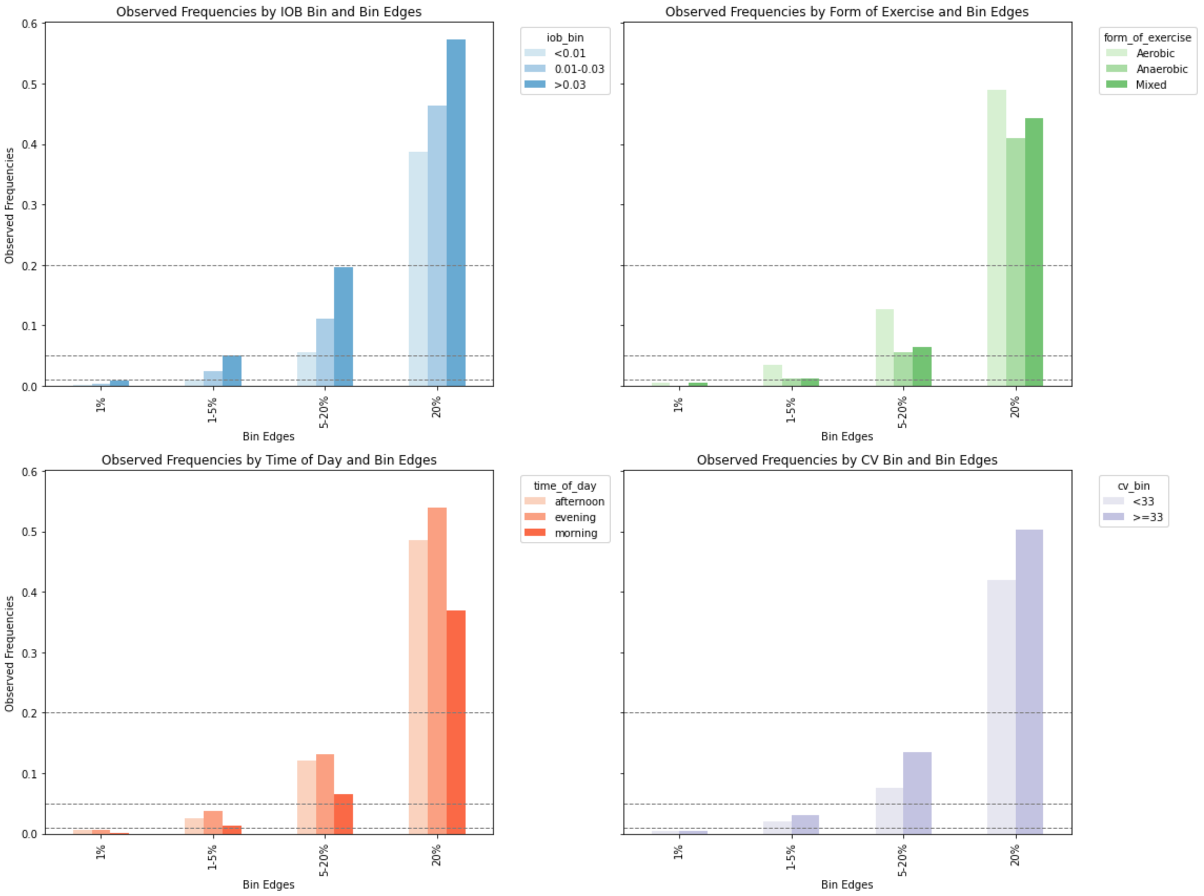


***Figure 6. Calibration of the GlucoseGo heatmap across key subgroups.***

*Bar plots show the observed rate of hypoglycaemia during exercise for each heatmap risk category (<5%, 5–20%, >20%), stratified by (A) insulin on board (IOB), (B) predominant form of exercise, (C) time of day, and (D) coefficient of variation (CV). For each subgroup, the observed rates fall within the intended risk thresholds indicated by the heatmap categories, confirming that the model's predictions remain well-calibrated across variations in IOB, exercise type, time of day, and glycaemic variability. Dashed horizontal lines indicate the thresholds for the heatmap risk categories: <5%, 5–20%, and >20%.*

### 8. SHAP Analysis: Variable Importance and Nonlinear Patterns

The complex XGBoost relied largely on clinically relevant, explainable variables, with only three *tsfresh* variables identified within the top 12 most important variables in the SHAP summary plot (Fig. 7). The eight most important, clinically relevant variables predicting risk of hypoglycaemia can be seen in the SHAP dependence plots in Fig. 8. Starting glucose (Fig. 8A) and exercise duration (Fig. 8B) have the greatest predictive power and show that lower starting glucose and longer durations will  increase the model's predicted risk of hypoglycemia. Higher predicted risk of hypoglycaemia also appears to be associated with higher insulin on board (Fig. 8C), aerobic exercise (Fig. 8D), negative glucose rate of change (Fig. 2H), and higher intensity exercise (Fig. 8F). Lower risk appears to be associated with exercising in the morning (Fig. 8E), and, a bit less clearly, higher HbA1c (Fig. 8G), more years since diagnosis  (Fig. 8I), and higher BMI  (Fig. 8J).

However, while these patterns do appear within the data, the model does not establish simple linear relationships. Many variables exhibit non-linear effects, where their contribution to the model's prediction varies across different values. For example, variables such as HbA1c, BMI, and years since diagnosis show highly non-linear patterns, making it difficult to extract a straightforward trend. Starting glucose, exercise duration, and IOB also display non-linear relationships, where their impact on predicted risk changes unevenly. This nonlinearity highlights the value of using a non-linear model, as it enables the capture of complex, nuanced patterns in the data that would be missed with a linear approach.

The difference in scale between the variables and thus the relative importance of these factors in prediction is very different: starting glucose and exercise duration have SHAP values around five times greater than the fourth and fifth most important predictors (time of day and type of exercise.

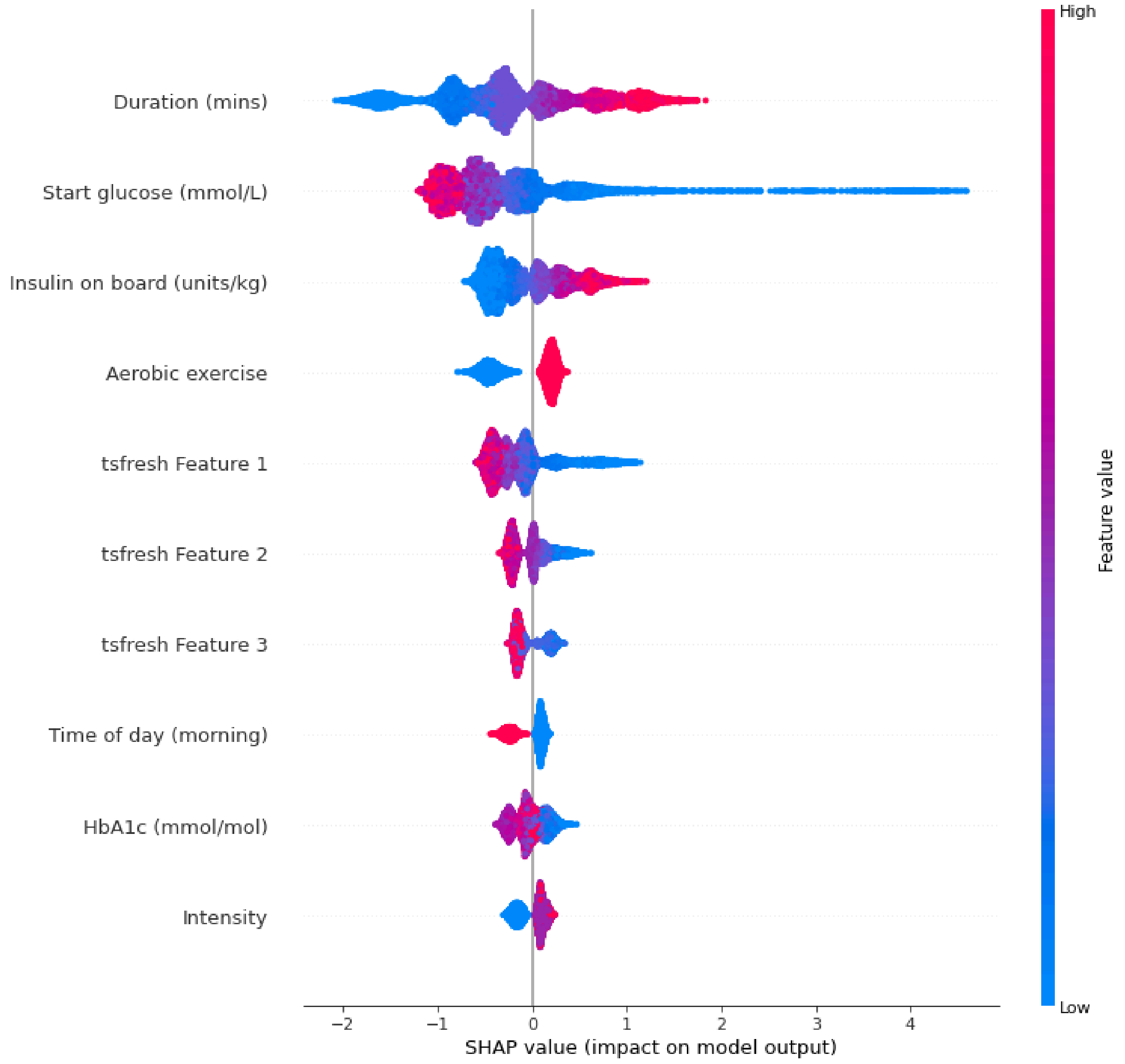


***Figure 7. SHAP Analysis for XGBoost Model with All Features.*** *SHAP values illustrate the contribution of each feature to the final prediction, shown as a shift in log-odds. Negative values indicate a decreased risk of hypoglycemia, and positive values an increased risk. Feature values are colour-coded from blue (low) to red (high). Variables beginning with "glc" are tsfresh features extracted from the one hour of CGM data prior to exercise. Features are ordered (top to bottom) by their overall average contribution to model predictions.*

*tsfresh Feature 1 represents the Continuous Wavelet Transform (CWT) coefficient. It computes the wavelet coefficients for various widths, specifically accessing the 11th coefficient. tsfresh Feature 2 computes the average value of the central second derivative, which is an approximation of the curvature or concavity of the time series. tsfresh Feature 3 extracts the angle (phase) of a coefficient from the Fast Fourier Transform (FFT) of the time series. tsfresh Feature 4, represents the partial autocorrelation excluding the contribution from the intermediate lags. tsfresh Feature 5 quantifies the approximate entropy statistic, used to measure the regularity and unpredictability of fluctuations in a time series. tsfresh Feature 6 calculates the ratio of energy for a specific chunk to the energy of the entire signal. tsfresh Feature 7 captures the imaginary component of a coefficient from the FFT of the time series. tsfresh Feature 8 extracts the angle (phase) of a coefficient from the FFT of the time series. tsfresh Feature 9 calculates the ratio of energy for a specific chunk to the energy of the entire signal. tsfresh Feature 10 represents the angle (phase) of a coefficient from the FFT of the time series. tsfresh Feature 11 captures the real component of a coefficient from the FFT of the time series.*

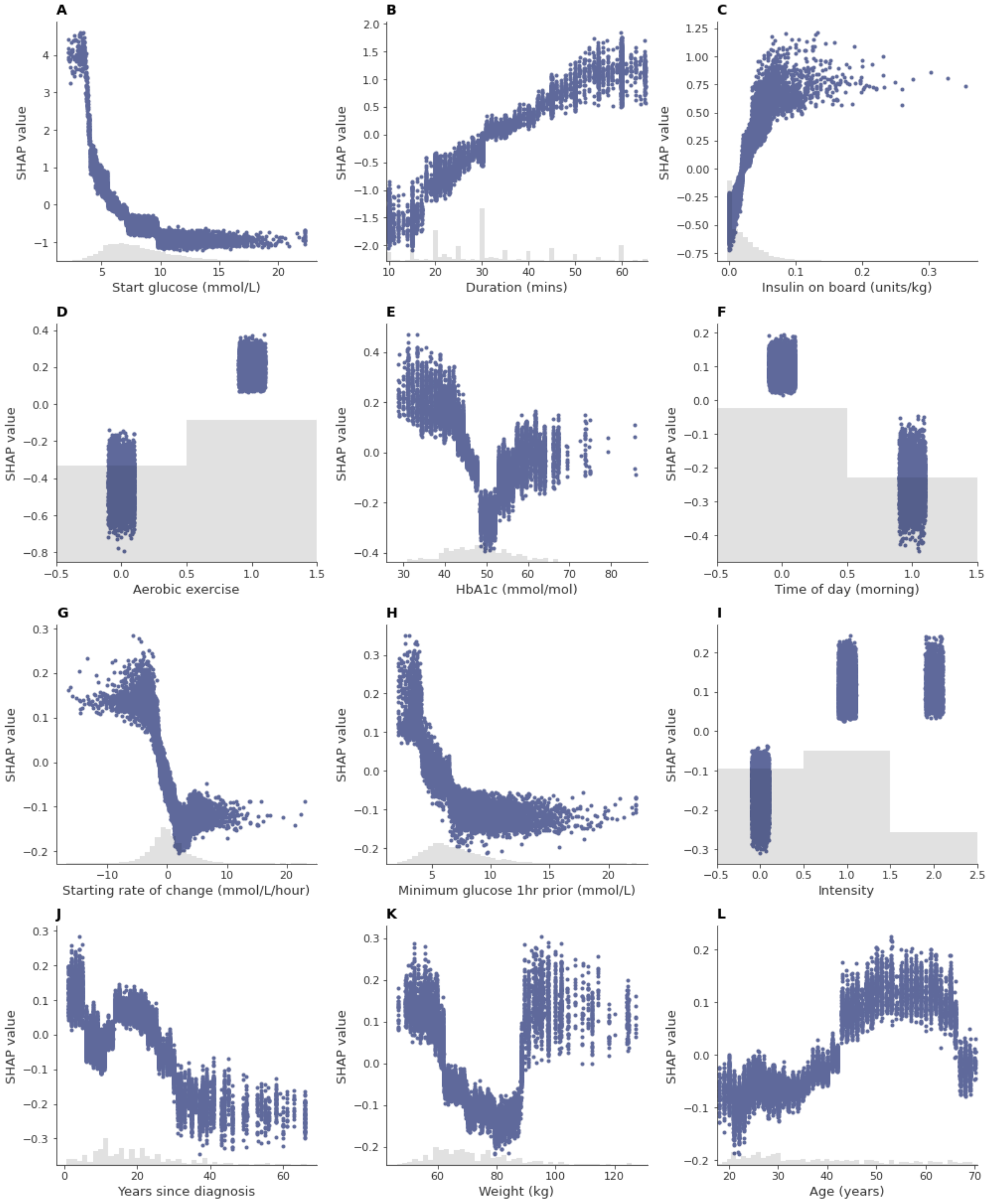


***Figure 8. SHAP dependence plots for clinically relevant features from the complex XGBoost model.*** *Blue points indicate the SHAP value (on the y-axis) for any given variable value (x-axis). SHAP values illustrate the contribution of each variable to the final prediction, shown as a shift in log-odds. Negative values indicate a decreased risk of hypoglycemia, and positive values an increased risk. The grey shaded area shows a histogram of variable values. The plot highlights the*

*six most clinically relevant variables with the greatest overall average contribution to model predictions.*

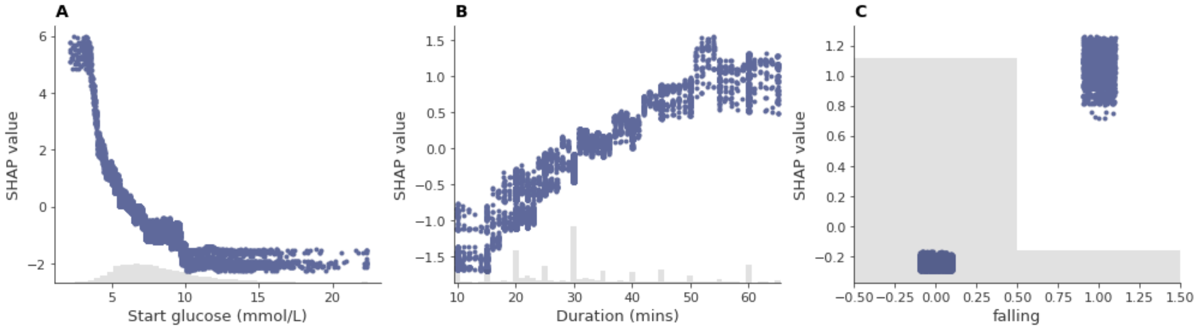


***Figure 9. SHAP dependence plots for clinically relevant features from the simplified XGBoost model.*** *Blue points indicate the SHAP value (on the y-axis) for any given variable value (x-axis). SHAP values illustrate the contribution of each variable to the final prediction, shown as a shift in log-odds. Negative values indicate a decreased risk of hypoglycemia, and positive values an increased risk. The grey shaded area shows a histogram of variable values. The plot highlights the three variables selected in feature selection. "Falling" is a binary variable that shows whether or not the rate of change was below 0.05 mmol/L / min (falling=1) or stable/rising (falling=0).*

### 9. Hyperparameters

| k-fold number | 1 | 2 | 3 | 4 | 5 | 6 | 7 | 8 | 9 | 10 |
|---|---|---|---|---|---|---|---|---|---|---|
| objective | binary:logistic | binary:logistic | binary:logistic | binary:logistic | binary:logistic | binary:logistic | binary:logistic | binary:logistic | binary:logistic | binary:logistic |
| base_score | 0.5 | 0.5 | 0.5 | 0.5 | 0.5 | 0.5 | 0.5 | 0.5 | 0.5 | 0.5 |
| booster | gbtree | gbtree | gbtree | gbtree | gbtree | gbtree | gbtree | gbtree | gbtree | gbtree |
| colsample_bylevel | 1 | 1 | 1 | 1 | 1 | 1 | 1 | 1 | 1 | 1 |
| colsample_bynode | 1 | 1 | 1 | 1 | 1 | 1 | 1 | 1 | 1 | 1 |
| colsample_bytree | 0.760 | 0.519 | 0.802 | 0.832 | 0.774 | 0.870 | 0.803 | 0.818 | 0.973 | 0.822 |
| gamma | 2 | 4 | 4 | 0 | 4 | 4 | 0 | 4 | 2 | 1 |
| learning_rate | 0.164 | 0.081 | 0.137 | 0.101 | 0.169 | 0.116 | 0.086 | 0.085 | 0.137 | 0.092 |
| max_delta_step | 0 | 0 | 0 | 0 | 0 | 0 | 0 | 0 | 0 | 0 |
| max_depth | 8 | 9 | 9 | 8 | 5 | 4 | 8 | 10 | 12 | 7 |
| min_child_weight | 8 | 7 | 9 | 7 | 8 | 1 | 8 | 5 | 7 | 8 |
| n_estimators | 236 | 588 | 976 | 312 | 7 | 282 | 233 | 219 | 389 | 908 |
| n_jobs | 12 | 12 | 12 | 12 | 12 | 12 | 12 | 12 | 12 | 12 |
| num_parallel_tree | 1 | 1 | 1 | 1 | 1 | 1 | 1 | 1 | 1 | 1 |
| predictor | auto | auto | auto | auto | auto | auto | auto | auto | auto | auto |
| random_state | 42 | 42 | 42 | 42 | 42 | 42 | 42 | 42 | 42 | 42 |
| reg_alpha | 4 | 3 | 1 | 2 | 1 | 2 | 1 | 1 | 3 | 1 |
| reg_lambda | 0 | 0 | 1 | 4 | 2 | 3 | 0 | 4 | 2 | 1 |
| scale_pos_weight | 1 | 1 | 1 | 1 | 1 | 1 | 1 | 1 | 1 | 1 |
| subsample | 0.866 | 0.871 | 0.734 | 0.692 | 0.993 | 0.997 | 0.933 | 0.991 | 0.801 | 0.860 |
| tree_method | exact | exact | exact | exact | exact | exact | exact | exact | exact | exact |
| validate_parameters | 1 | 1 | 1 | 1 | 1 | 1 | 1 | 1 | 1 | 1 |
| eta | 0.234 | 0.157 | 0.244 | 0.131 | 0.211 | 0.155 | 0.163 | 0.219 | 0.154 | 0.161 |
| seed | 42 | 42 | 42 | 42 | 42 | 42 | 42 | 42 | 42 | 42 |

***Table 2. Tuned hyperparameter for the 10 k-fold models for the complex XGBoost.***

| k-fold number | 1 | 2 | 3 | 4 | 5 | 6 | 7 | 8 | 9 | 10 |
|---|---|---|---|---|---|---|---|---|---|---|
| objective | binary:logistic | binary:logistic | binary:logistic | binary:logistic | binary:logistic | binary:logistic | binary:logistic | binary:logistic | binary:logistic | binary:logistic |
| base_score | 0.5 | 0.5 | 0.5 | 0.5 | 0.5 | 0.5 | 0.5 | 0.5 | 0.5 | 0.5 |
| booster | gbtree | gbtree | gbtree | gbtree | gbtree | gbtree | gbtree | gbtree | gbtree | gbtree |
| colsample_bylevel | 1 | 1 | 1 | 1 | 1 | 1 | 1 | 1 | 1 | 1 |
| colsample_bynode | 1 | 1 | 1 | 1 | 1 | 1 | 1 | 1 | 1 | 1 |
| colsample_bytree | 0.99 | 0.77 | 0.94 | 0.89 | 0.88 | 0.69 | 0.83 | 0.82 | 0.72 | 0.89 |
| enable_categorical | FALSE | FALSE | FALSE | FALSE | FALSE | FALSE | FALSE | FALSE | FALSE | FALSE |
| gamma | 4 | 4 | 4 | 3 | 0 | 4 | 5 | 5 | 4 | 1 |
| gpu_id | -1 | -1 | -1 | -1 | -1 | -1 | -1 | -1 | -1 | -1 |
| learning_rate | 0.48 | 0.19 | 0.36 | 0.11 | 0.27 | 0.24 | 0.24 | 0.31 | 0.15 | 0.22 |
| max_delta_step | 0 | 0 | 0 | 0 | 0 | 0 | 0 | 0 | 0 | 0 |
| max_depth | 8 | 5 | 8 | 3 | 5 | 3 | 9 | 4 | 11 | 3 |
| min_child_weight | 5 | 7 | 5 | 1 | 10 | 2 | 8 | 10 | 7 | 1 |
| n_estimators | 325 | 867 | 586 | 637 | 489 | 135 | 49 | 322 | 584 | 507 |
| n_jobs | 12 | 12 | 12 | 12 | 12 | 12 | 12 | 12 | 12 | 12 |
| num_parallel_tree | 1 | 1 | 1 | 1 | 1 | 1 | 1 | 1 | 1 | 1 |
| predictor | auto | auto | auto | auto | auto | auto | auto | auto | auto | auto |
| random_state | 42 | 42 | 42 | 42 | 42 | 42 | 42 | 42 | 42 | 42 |
| reg_alpha | 5 | 1 | 0 | 4 | 2 | 1 | 0 | 1 | 1 | 2 |
| reg_lambda | 3 | 3 | 5 | 3 | 5 | 5 | 1 | 4 | 3 | 1 |
| scale_pos_weight | 1 | 1 | 1 | 1 | 1 | 1 | 1 | 1 | 1 | 1 |
| subsample | 0.832 | 0.905 | 0.920 | 0.959 | 0.582 | 0.699 | 0.696 | 0.893 | 0.602 | 0.779 |
| tree_method | exact | exact | exact | exact | exact | exact | exact | exact | exact | exact |
| validate_parameters | 1 | 1 | 1 | 1 | 1 | 1 | 1 | 1 | 1 | 1 |
| eta | 0.142 | 0.262 | 0.117 | 0.191 | 0.263 | 0.155 | 0.254 | 0.117 | 0.114 | 0.128 |
| seed | 42 | 42 | 42 | 42 | 42 | 42 | 42 | 42 | 42 | 42 |

***Table 3. Tuned hyperparameter for the 10 k-fold models for the simplified XGBoost.***

### 10. Learning Curves

A

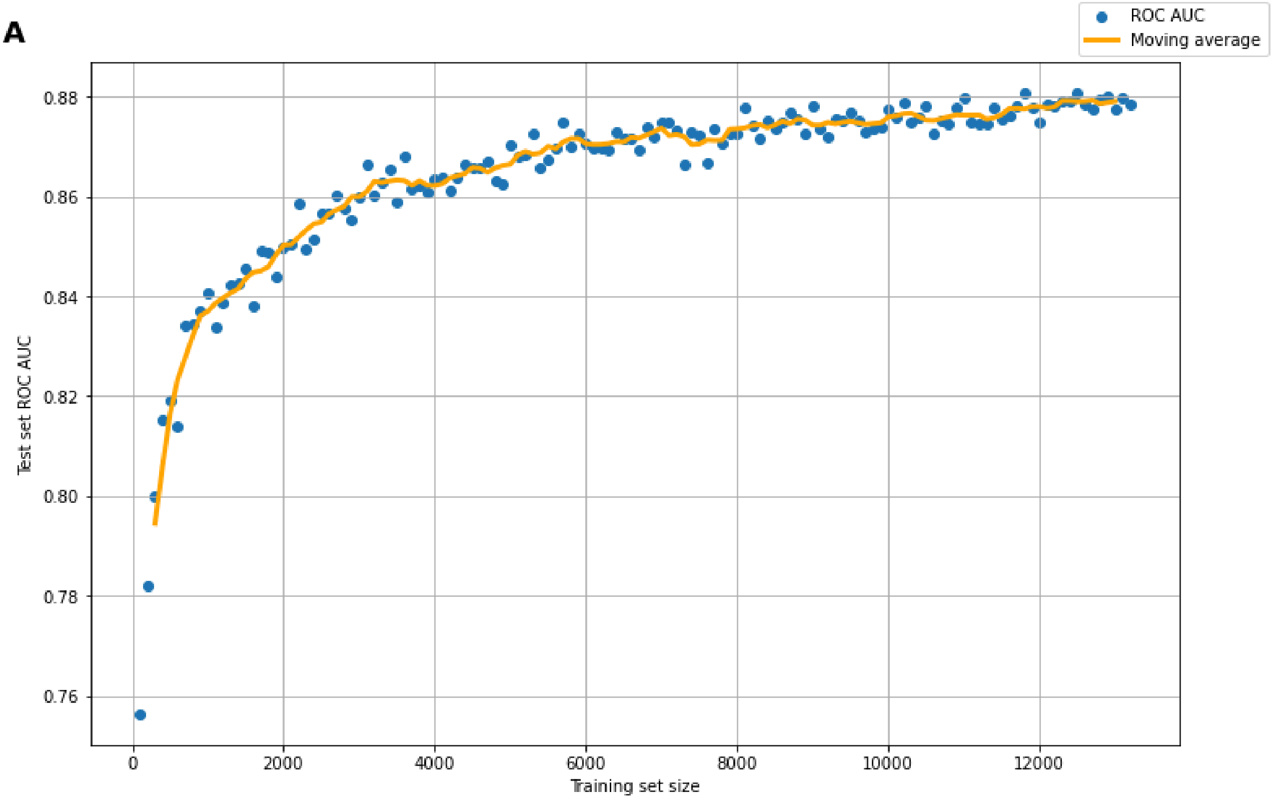


B

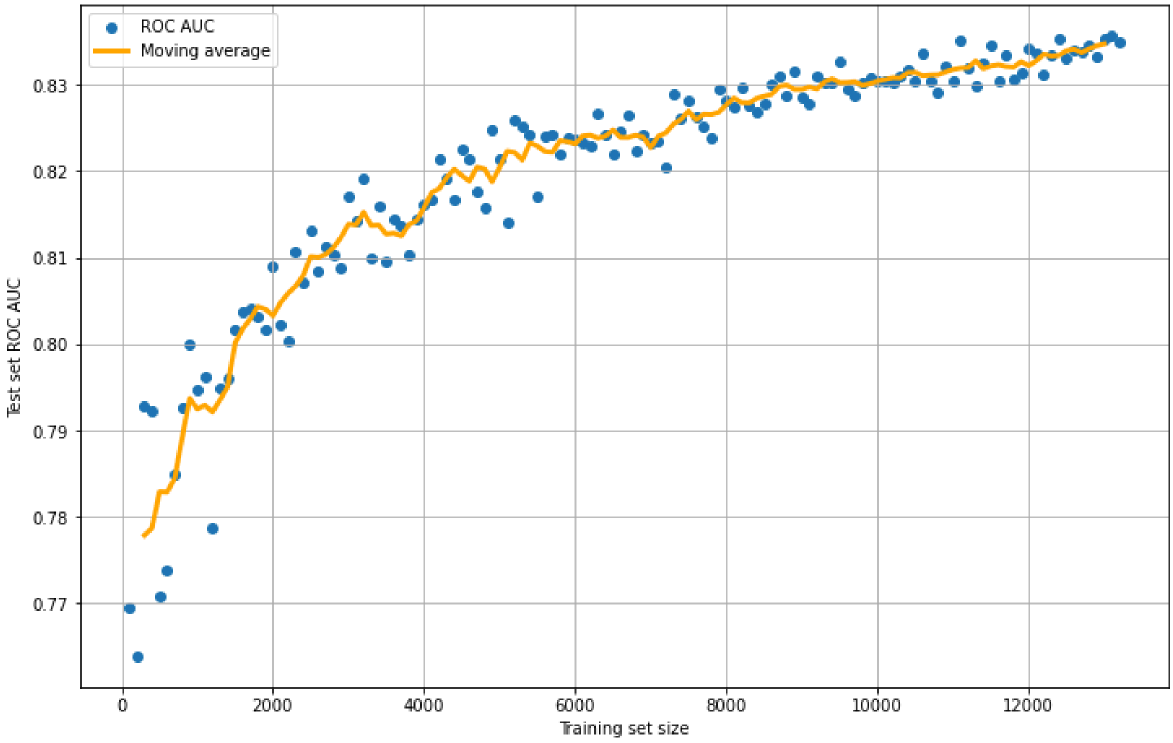


***Figure 10. Learning curves showing performance of complex and simplified XGBoost with increasing data.*** *Learning curves depict a model's performance as it trains on increasing amounts of data, helping determine if more data improves its accuracy. Blue dots represent the model's test performance at different training sizes, with each iteration increasing by 100 data points. The*

*yellow line shows the moving average of the performance over the last five data points, providing a smoothed trend. Testing is consistently done on 20% of the available data for each iteration. A: the curve for the full-featured XGBoost. B: the two-featured XGBoost, with start glucose and duration only.*
